# Foundation models capture human and environmental context to improve dengue forecasting

**DOI:** 10.64898/2026.11.05.26367335

**Authors:** John-Stuart Brittain, Cathal Mills, Ciara Judge, Leonardo S. Bastos, Laís Picinini Freitas, Luiz Max Carvalho, Eduardo Correa Araujo, Flávio C. Coelho, Yanhua Xu, Benjamin Reddy, Rama Kumar Pasumarthi, Tomer Shekel, Shravya Shetty, Amy Chung-Yu Chou, Anna Carter, Arbaaz Muslim, Gautam Prasad, Kasumi Widner, Monica Bharel, Samuel V. Scarpino, Seth Flaxman, Samir Bhatt, Joseph L.H. Tsui, Moritz U.G. Kraemer

**Author notes:** Correspondence should be addressed to John-Stuart Brittain and Moritz U.G. Kraemer. These authors contributed equally (John-Stuart Brittain, Cathal Mills).

## Abstract

Accurate and timely probabilistic dengue forecasts can support public health preparedness, but producing reliable forecasts across locations and time horizons remains challenging. We evaluated five statistical, machine-learning, deep-learning, and foundation-model approaches for forecasting monthly reported dengue cases across 5,570 Brazilian municipalities at 1-, 3-, 6- and 12-month horizons, with forecasts backtested from January 2020 to August 2025. The time-series foundation model TimesFM maintained comparatively stable probabilistic forecast performance at longer horizons, while most other models showed declining performance. We then tested whether spatial representations of human and environmental context from a Population Dynamics Foundation Model (PDFM) could improve forecasts through post-hoc residual adjustment. PDFM adjustment improved mean probabilistic forecast performance, measured by the Weighted Interval Score (WIS), in 19 of 20 model-horizon combinations, although the magnitude and frequency of improvements varied across models, horizons and municipalities. The largest improvements were observed for N-BEATS, including a reduction in mean WIS of 1.13 at the six-month horizon (95% CI [0.84, 1.48]; Diebold–Mariano p < 0.0001), with improvements also observed for XGBoost. The proportion of forecasts improved by PDFM generally increased with locally reported dengue case counts, although this relationship varied between models. Covariate selection, spatial fitting and forecast aggregation strategies showed strong model-dependent effects, while monthly model retraining provided no consistent advantage in the settings evaluated. Together, these findings highlight the potential of pretrained representations for dengue forecasting, with benefits dependent on forecasting architecture, horizon and modelling configuration.

## Introduction

Dengue is a major vector-borne disease, causing substantial annual morbidity and mortality as well as economic and social disruption. Dengue virus (DENV) is transmitted primarily by *Aedes aegypti* and, to a lesser degree, *Ae. albopictus* in tropical and temperate regions, with transmission intensity shaped by climatic, environmental, socioeconomic, demographic and biological factors, including population immunity and circulating DENV serotypes (1). The relative importance of these factors varies over time and by location and is highly context-specific (2). The number of reported dengue cases reached record levels in 2024, with more than 14 million cases reported worldwide. In Brazil alone, more than 10 million dengue cases were reported through surveillance notifications, alongside over 6,000 dengue-related deaths (3). The record epidemic in Brazil and the Americas coincided with *El Niño*-associated temperature anomalies and substantial southward geographical expansion of dengue transmission into previously less-affected temperate regions (4). Geographical expansion and increased cases in endemic areas highlight the vulnerability of cities in the Americas to climate-related shifts in temperature and precipitation, creating new challenges for dengue preparedness and response.

Anticipating where and when reported dengue cases will increase, and by how much, is therefore important for targeting preparedness and response. Probabilistic forecasts characterise uncertainty in these future outcomes rather than providing a single expected trajectory, and can support decisions about where and when public health resources are needed. Forecasts over longer time horizons, from several months to a year ahead, can inform advance planning and resource allocation across regions, while shorter-term forecasts can support more immediate decisions around healthcare capacity, risk communication and targeted vector-control interventions. In Brazil, dengue preparedness activities routinely include intensified vector surveillance and campaigns to reduce mosquito breeding sites before seasonal peaks, alongside emergency healthcare mobilisation and redistribution of clinical resources during major epidemics (5–7).

Current approaches to dengue forecasting have primarily relied on historical incidence alongside climatic, demographic and, in some cases, immunological predictors. Existing approaches include spatiotemporal models incorporating climatic predictors (8–10), probabilistic models for seasonal epidemic forecasting (11), and ensemble methods combining predictions across multiple models (12–14). For example, the 2024 edition of the Infodengue-Mosqlimate Dengue Challenge (IMDC24) evaluated probabilistic forecasts from seven statistical and machine-learning models across five Brazilian states, demonstrating substantial variation in performance across models, locations and epidemic seasons, and highlighting the potential value of ensemble approaches (15). However, local population dynamics that shape transmission risk, including human behaviour, mobility, and interactions with the social and built environment, remain difficult to capture fully using conventional epidemiological and climatic covariates (16). The increasing availability of digital data provides new opportunities to characterise these dynamics, including through aggregated signals of mobility, behaviour and population activity. Population Dynamics Foundation Model (PDFM) provides one scalable approach to integrating these heterogeneous factors, learning transferable spatial representations from multimodal information including aggregated search trends, mobility patterns, built-environment characteristics, weather and air quality (17). In Brazil, PDFM embeddings have recently shown utility for subnational population estimation, including improvements over traditional hand-crafted geospatial covariates in some settings (18). Whether these representations improve predictive performance beyond that achieved by existing infectious disease forecasting models using conventional epidemiological and environmental covariates remains, to our knowledge, unknown.

Beyond representing spatial and contextual information, foundation models can also learn representations of temporal dynamics through large-scale pretraining (19). Time-series foundation models are trained on large collections of time-series data spanning diverse domains, including those unrelated to infectious diseases, with the aim of learning representations that generalise across downstream forecasting tasks (20, 21). Some can be adapted or fine-tuned using task-specific data to further improve performance (22). Recent studies have applied time-series foundation models to infectious-disease forecasting, including dengue and other communicable diseases (12, 23). Automated model development using multimodal data and foundation-model embeddings has also shown promise across geospatial prediction tasks, including infectious disease modelling (24).

Here, we evaluate whether pretrained temporal and spatial representations can strengthen forecasting of dengue epidemic dynamics, measured by monthly reported cases at 1-, 3-, 6- and 12-month horizons. The analysis encompasses 5,570 municipality-level administrative units across Brazil, including the Federal District and Fernando de Noronha (hereafter collectively referred to as municipalities). We test two complementary applications of foundation models: a time-series foundation model pretrained on large-scale temporal data, and spatial embeddings from PDFM representing human and environmental characteristics. We compare five forecasting models spanning statistical, machine-learning, deep-learning and foundation-model paradigms, and assess whether using PDFM embeddings improves probabilistic forecast performance beyond that achieved using historical incidence and conventional covariates. We further assess how covariate selection, spatial fitting strategy, forecast aggregation and retraining strategy influence forecasting performance. Finally, we use Empirical Research Assistance (ERA), an Artificial Intelligence (AI) system combining large language models and tree search to iteratively develop scientific software (25), to test whether PDFM embeddings are independently selected for dengue forecasting. ERA was provided with the same epidemiological, climatic and PDFM inputs as the primary forecasting models, alongside spatial information.

## Results

Reported dengue case counts varied substantially across Brazilian municipalities and through time, with pronounced epidemic peaks between January 2010 and August 2025 (Figure 1A–B). We generated and evaluated probabilistic dengue forecasts across 5,570 Brazilian municipalities using five forecasting models spanning statistical (SARIMA), conventional machine-learning (XGBoost), deep-learning (TCN and N-BEATS) and foundation-model (TimesFM) approaches (see Methods). Four of the models were trained using data from January 2010 to December 2019, while TimesFM was applied without fitting or fine-tuning its pretrained model parameters to the study data; forecasts were evaluated retrospectively over a common target-month window from January 2020 to August 2025 at 1-, 3-, 6- and 12-month horizons (Figure 1B). To represent local human and environmental context, we used 330-dimensional municipality-level PDFM embeddings from a single October 2023 snapshot, comprising components derived from Search Trends, Maps & Busyness, and Weather & Air Quality (Figure 1C). Correlations among embedding dimensions indicate shared structure within the learned representation, while representative dimensions demonstrate spatial correspondence with nighttime light intensity (Pearson r = 0.51), population count (r = -0.23) and temperature (r = 0.78; Figure 1C–D). Forecast performance was assessed primarily using the Weighted Interval Score (WIS), calculated on the log(1 + cases) scale to facilitate comparison across municipalities and time, with lower values indicating better probabilistic forecast performance (Methods; Figure 1E). The resulting predictive distributions quantify uncertainty across forecast horizons, as illustrated for São Paulo state using example TCN forecasts at 1-, 3-, 6- and 12-month horizons from a common December 2019 forecast origin (Figure 1F). We first compared the forecasting models without PDFM adjustment, before testing whether municipality-level PDFM embeddings provided complementary information through residual adjustment.

**Figure 1.**
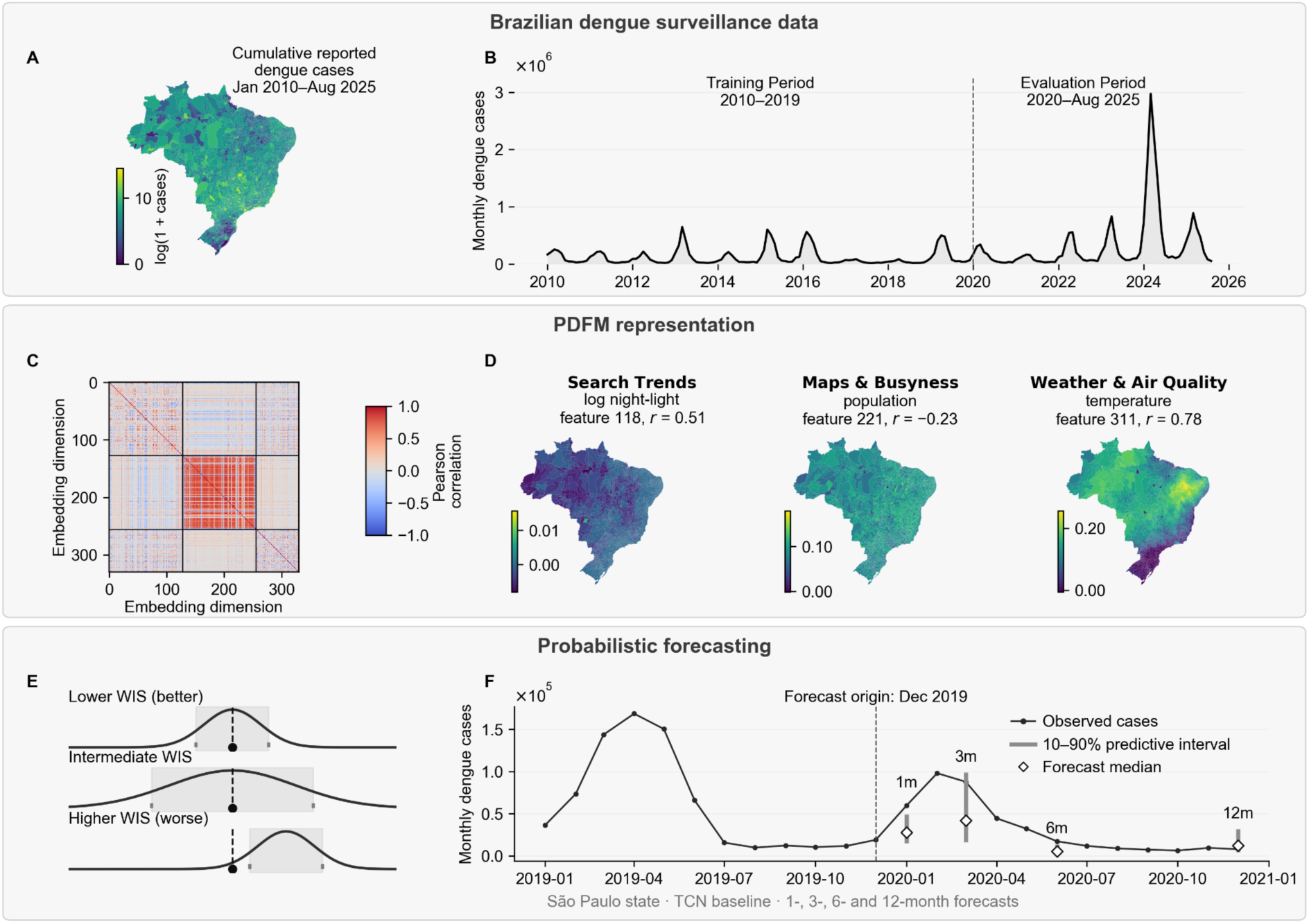
Study setting, Population Dynamics Foundation Model (PDFM) representations and probabilistic forecasting framework. (A) Municipality-level cumulative reported dengue cases across Brazil from January 2010 to August 2025, shown on a log(1 + cases) scale. (B) National monthly reported dengue cases from January 2010 to August 2025, showing the January 2010–December 2019 model-training period and January 2020–August 2025 forecast-evaluation period. All values shown are observed case counts. (C) Pairwise Pearson correlation matrix of the 330-dimensional municipality-level PDFM embeddings. Black lines delineate the three modality-specific components of the representation: aggregated Search Trends (dimensions 0–127), Maps & Busyness (dimensions 128–255), and Weather & Air Quality (dimensions 256–329). (D) Spatial distributions of representative embedding dimensions from each PDFM component. Within each component, the displayed dimension was selected as that with the strongest absolute municipality-level correlation with an external spatial characteristic: log-transformed nighttime light intensity for Search Trends, population for Maps & Busyness, and temperature for Weather & Air Quality; the corresponding Pearson correlation coefficient (r) is shown above each map. (E) Schematic examples illustrating how the Weighted Interval Score (WIS) evaluates probabilistic forecasts. Lower WIS indicates better probabilistic forecast performance, reflecting sharper predictive distributions that remain well calibrated; higher WIS reflects wider predictive distributions and/or failure to cover the observed outcome. (F) Illustration of probabilistic forecasts at different horizons, showing observed state-level dengue cases for São Paulo from January 2019 to December 2020 and TCN forecasts fitted to the aggregated state-level case series from a common December 2019 forecast origin for target months January, March, June and December 2020 (1-, 3-, 6- and 12-month horizons, respectively). Vertical bars show the 10–90% predictive interval and diamonds show the forecast median.

### TimesFM maintains strong relative performance at longer forecast horizons

For São Paulo, the municipality which had the highest cumulative number of reported dengue cases among Brazilian municipalities over the study period, the five forecasting models produced distinct predictive distributions (Figure 2A). Across all municipalities, probabilistic forecast performance varied by model and forecast horizon (Figure 2B; Supplementary Table S1). At the 1-month horizon, TCN achieved the lowest mean WIS (1.27), followed by XGBoost (1.38), while SARIMA and TimesFM performed similarly (mean WIS 1.46). As forecast horizon increased, performance deteriorated for most models, while TimesFM remained comparatively stable, achieving mean WIS of 1.56, 1.53 and 1.51 at 3-, 6- and 12-month horizons, respectively. TCN was the next-best performing model at these horizons, with mean WIS of 1.89, 2.08 and 2.16, respectively. Mean WIS was calculated across all municipality-month forecasts within each model–horizon combination (n = 378,760), with each observation contributing equally. WIS distributions were right-skewed, with means exceeding medians across models and horizons; for example, at 12 months, N-BEATS had a mean WIS of 3.53 compared with a median of 1.07, while TimesFM had a mean of 1.51 and median of 0.94.

**Figure 2.**
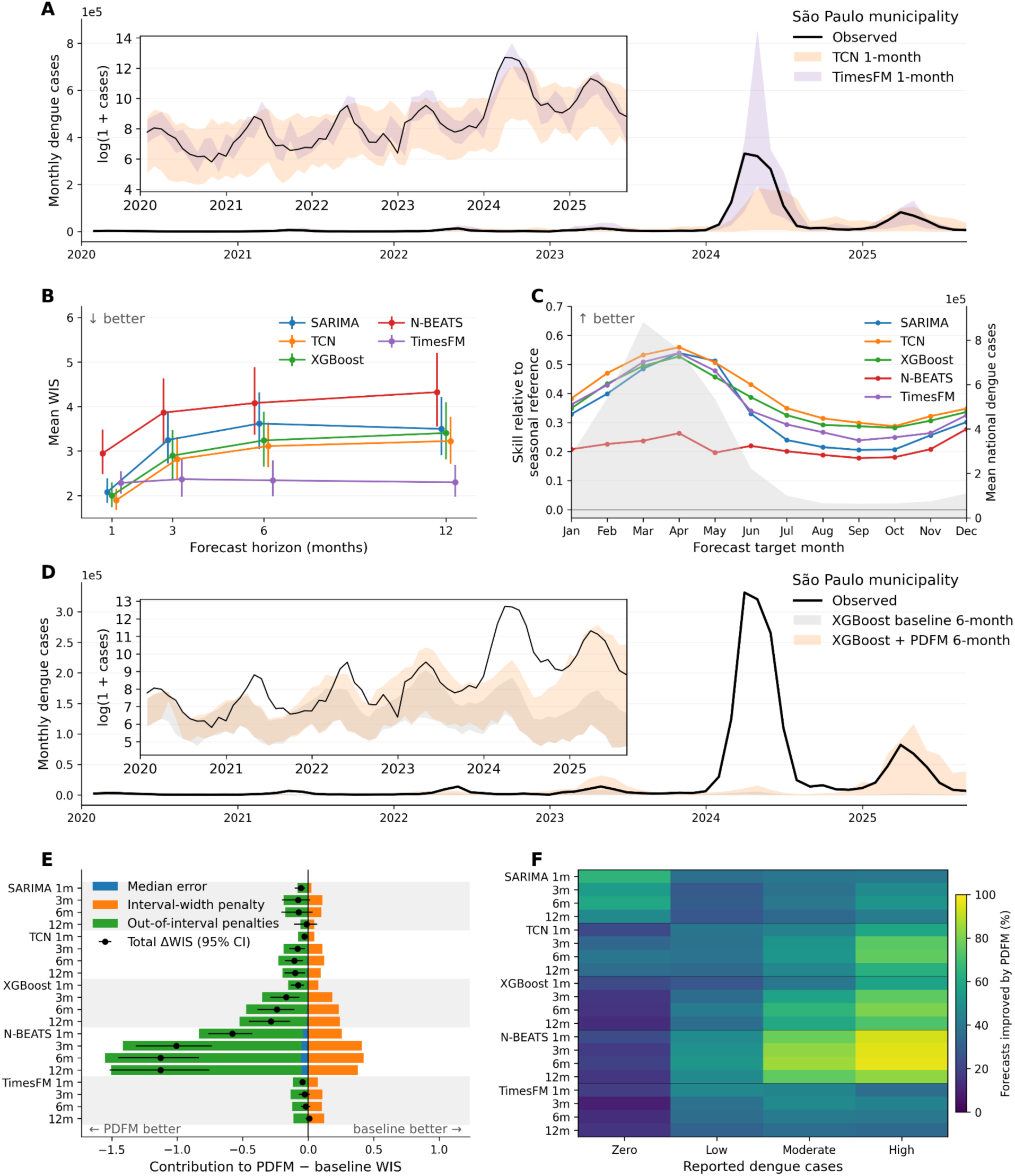
Baseline dengue forecasting performance and effects of Population Dynamics Foundation Model (PDFM) residual adjustment. Forecasts were evaluated over a common target-month window from January 2020 to August 2025 across Brazilian municipalities. (A) Observed monthly dengue cases and representative one-month-ahead probabilistic forecasts from a temporal convolutional network (TCN) and TimesFM for São Paulo municipality (in the state of São Paulo), selected as the municipality with the highest cumulative number of reported dengue cases over the study period. Shaded regions show the 10– 90% predictive intervals; the inset shows the same forecasts and observations on the log(1 + cases) scale. (B) Mean Weighted Interval Score (WIS) across municipality-month forecasts for each model and forecast horizon. Points show mean WIS and vertical bars show 95% confidence intervals estimated using a temporal moving-block bootstrap (3-month blocks; 2,000 replicates); lower WIS indicates better probabilistic forecast performance. SARIMA, Seasonal Autoregressive Integrated Moving Average; TCN, Temporal Convolutional Network; XGBoost, extreme gradient boosting; N-BEATS, Neural Basis Expansion Analysis for Interpretable Time Series. (C) Seasonal variation in one-month log-relative WIS skill compared with a municipality-specific seasonal reference forecast. The seasonal reference forecast was constructed separately for each municipality and calendar month using the 2010–2019 training data (see Methods). Skill is defined as log(1 + WIS_seasonal_) − log(1 + WIS_model_), such that positive values indicate better performance than the seasonal reference forecast. Lines show mean skill across municipalities by calendar month; grey shading shows mean national reported dengue cases in the corresponding calendar month during the evaluation period. (D) Six-month-ahead XGBoost forecasts for the same municipality before and after PDFM residual adjustment. Shaded regions show the 10–90% predictive intervals; the inset shows the same forecasts and observations on the log(1 + cases) scale. (E) Decomposition of the mean change in WIS following PDFM adjustment (ΔWIS = WIS_PDFM_ − WIS_base_) into contributions from median forecast error, predictive-interval width and out-of-interval penalties (see Methods). Negative contributions improve WIS and positive contributions increase WIS. Black points show the sum of these contributions, corresponding to total ΔWIS, with horizontal bars showing 95% temporal moving-block-bootstrap confidence intervals. (F) Percentage of municipality-month forecasts improved by PDFM adjustment (ΔWIS < 0), stratified retrospectively by reported dengue case counts in the forecast target month. Municipality-months with no reported cases form the zero-case stratum; within each target month and model-horizon combination, observations with positive reported case counts were divided into low, moderate and high groups using the 33rd and 67th percentiles of their case-count distribution.

To examine how forecasting performance varied through time, we compared one-month forecasts against a municipality-specific seasonal reference forecast (Figure 2C). All models showed positive mean skill relative to this reference in every calendar month, although the magnitude of this advantage varied seasonally. For each model and calendar month, mean skill was calculated across municipalities and evaluation years (2020–2025). Relative skill was generally greatest during the early months of the year, when reported dengue case counts were also higher, and declined later in the year.

We also examined geographical variation in forecasting performance across Brazil’s five macroregions (North, Northeast, Southeast, South and Central-West). The model rankings were consistent across macroregions, with TCN achieving the lowest mean WIS at the one-month horizon and TimesFM at 3-, 6- and 12-month horizons (Supplementary Table S2). However, the magnitude of these differences varied geographically. At one month, TCN’s mean WIS advantage over TimesFM was substantially smaller in the South (ΔWIS = −0.078, TCN minus TimesFM) than in the other macroregions (−0.190 to −0.223). This difference was concentrated in Rio Grande do Sul (ΔWIS = −0.003) and Santa Catarina (+0.005), whereas Paraná showed a clearer TCN advantage (−0.232). TCN outperformed TimesFM in 68.4% and 65.4% of municipalities in Rio Grande do Sul and Santa Catarina, respectively, compared with 92.2% in Paraná. At longer horizons, TimesFM retained the lowest mean WIS across all macroregions, with larger advantages over TCN in the Central-West, Southeast and South than in the North and Northeast. These findings demonstrate that the relative performance of forecasting architectures varies geographically, even where the leading model remains consistent at macroregional level.

### PDFM embeddings provide complementary predictive information

We next tested whether municipality-level PDFM embeddings provided predictive information complementary to the baseline forecasting models using residual adjustment (Methods). For example, at the six-month horizon in the municipality of São Paulo, PDFM adjustment altered the XGBoost predictive distribution, including during the major 2024 epidemic (Figure 2D). Averaged across all municipality-month forecasts, PDFM adjustment reduced mean WIS for 19 of 20 model– horizon combinations, although the magnitude and consistency of improvement varied substantially between models and horizons (Supplementary Table S3). Improvements were largest for N-BEATS, for which mean ΔWIS (PDFM − base) ranged from −0.58 at one month to −1.13 at six and 12 months horizons, and XGBoost, for which improvements increased from −0.08 at one month to −0.29 at 12 months. For N-BEATS and XGBoost, 95% temporal bootstrap confidence intervals for mean ΔWIS excluded zero at all four horizons. Following Bonferroni correction across the 20 model–horizon comparisons, Diebold–Mariano tests supported improved predictive accuracy at all four horizons for N-BEATS and at the one-month horizon for XGBoost (Supplementary Table S3). TCN models showed improvements at 1, 3 and 6 months supported by temporal bootstrap confidence intervals, although only the one-month comparison remained statistically significant after Bonferroni correction of the Diebold–Mariano tests. At a 12-month horizon, the temporal bootstrap indicated improvement (mean ΔWIS = −0.10, 95% CI [−0.18, −0.03]), but the Diebold–Mariano test did not reject equal predictive accuracy (p = 0.14) and bootstrap inference was sensitive to block length (Supplementary Table S3). Evidence of benefit at 12 months was therefore inconclusive. For SARIMA and TimesFM, evidence of benefit was limited to the one-month horizon; at longer horizons, bootstrap confidence intervals spanned zero and Diebold–Mariano tests did not support improved predictive accuracy. PDFM effects also varied through time at longer forecast horizons (Supplementary Figures S2–S3).

Decomposition of ΔWIS showed that the largest PDFM improvements arose primarily from reductions in out-of-interval penalties, which outweighed increases in predictive-interval width, while changes in median forecast error were comparatively small (Figure 2E). This pattern was particularly pronounced for N-BEATS and XGBoost at longer horizons.

The effect of PDFM adjustment also varied with the number of locally reported dengue cases in the forecast target month (Figure 2F). Because absolute differences in WIS can increase with forecast-error magnitude, we examined the proportion of individual municipality-month forecasts improved by PDFM within each case-count stratum. The proportion improved generally increased with reported case counts for several models; for example, at the six-month horizon, PDFM adjustment reduced WIS for 96.2% of N-BEATS forecasts in the highest case-count stratum, with a mean ΔWIS of −5.05 (95% CI [−5.52, −4.66]). The corresponding mean ΔWIS was −0.19 (95% CI [−0.32, −0.09]) in the lowest positive case-count stratum, with a high-minus-low difference of −4.87 (95% CI [−5.21, −4.54]; Supplementary Table S4). Conversely, PDFM adjustment worsened performance for zero-case observations in several models, including N-BEATS at six months (mean ΔWIS = +0.36, 95% CI [0.30, 0.41]). These exploratory findings indicate that the magnitude and direction of PDFM effects vary with observed case counts, with substantial differences between forecasting models.

The benefits of PDFM residual adjustment also varied geographically, with patterns differing across forecasting architectures (Supplementary Table S5). At the 12-month horizon, TCN and XGBoost showed particularly large absolute reductions in WIS in the South (mean ΔWIS = −0.306 and −0.524, respectively). However, for TCN, these larger absolute improvements did not translate into significantly greater log-relative skill than in the Northeast or Southeast. In contrast, N-BEATS showed consistently greater relative benefits in the Southeast and South than in the Northeast across all four horizons, while XGBoost showed greater relative benefits in the Central-West than in the Northeast at longer horizons. These findings indicate that the geographical distribution of PDFM benefits depends on the forecasting architecture and the measure of improvement considered, rather than following a consistent regional pattern across models.

### Forecasting performance depends on modelling strategy

We next examined whether forecasting models benefited consistently from increasing the number of epidemiological and environmental covariates included. We compared the full covariate specification with a minimal set comprising log(1 + cases) at lag 1, the mean of minimum temperature over lags 1–2, and the mean of precipitation over lags 1–2. Models additionally retained their architecture-specific historical dengue case inputs or autoregressive structure (Table 2). The effect of covariate complexity was strongly model-dependent (Figure 3A; Supplementary Table S6). N-BEATS and SARIMA consistently benefited from the minimal covariate set, which yielded lower mean WIS, averaged across evaluation months within each municipality, in 79–87% of municipalities for N-BEATS and >97% for SARIMA across forecast horizons. TCN showed the opposite pattern, with the full covariate set producing better forecasts across horizons and approximately 73–76% of municipalities favouring the full configuration. XGBoost showed a horizon-dependent transition: the minimal covariate set modestly improved performance at one month, produced little difference at three months, while the full covariate set performed better at six and 12 months (Figure 3A; Supplementary Table S6). TimesFM was comparatively insensitive to covariate inclusion, with little difference between the full and minimal covariate sets across forecast horizons (Figure 3A; Supplementary Table S6). Overall, increasing predictor complexity did not provide a consistent forecasting advantage: the value of additional covariates depended on the model and, in some cases, forecast horizon.

**Figure 3.**
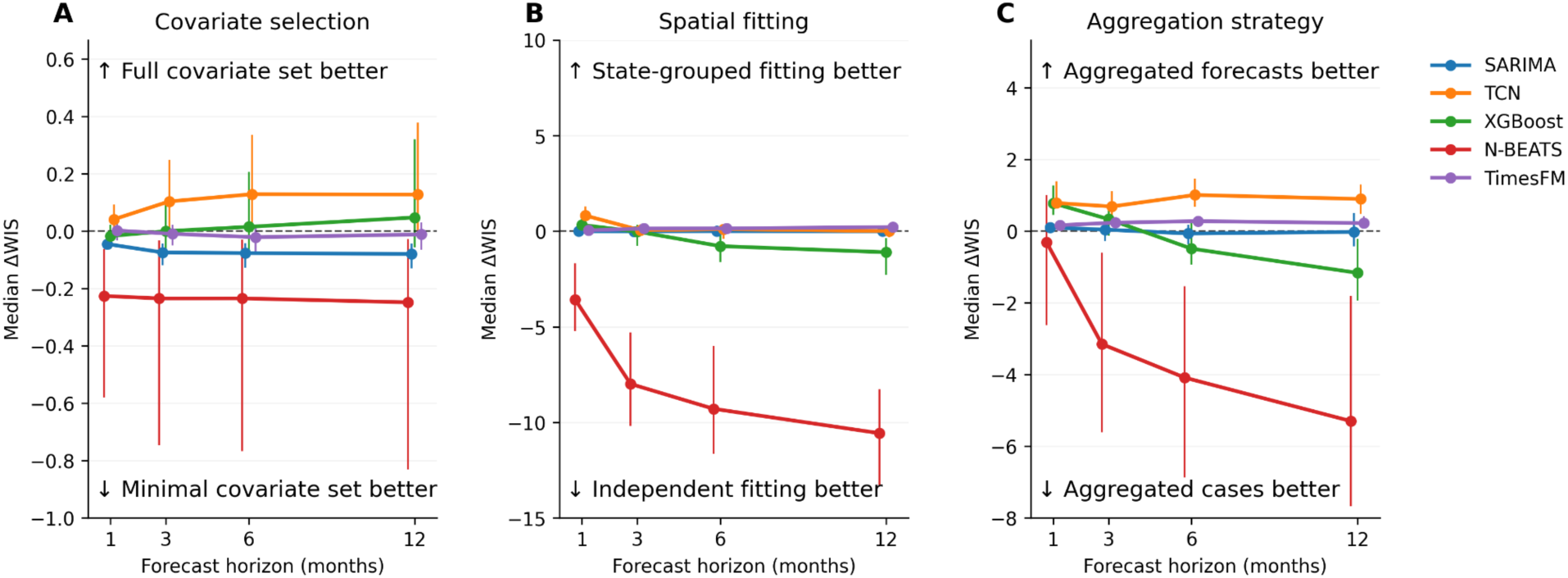
Forecast configuration effects depend on the forecasting model. Points show the median change in Weighted Interval Score (ΔWIS) across spatial units, with vertical bars indicating the interquartile range (25th–75th percentiles). Paired forecast differences were averaged over the January 2020–August 2025 evaluation period within each municipality or state before summarising across spatial units. (A) Difference in forecast performance between models using minimal and full covariate sets (see Table 1) across 5,570 municipalities. Negative values indicate better performance with the minimal covariate set. (B) Comparison of municipality-level forecasts from models fitted independently to each municipality versus models fitted jointly across municipalities within each state, evaluated for the same 20 municipalities with the highest cumulative reported dengue case counts over the study period. Negative ΔWIS values favour independent municipality-level fitting. (C) Comparison of forecasts fitted directly to aggregated state-level case series with state-level forecasts obtained by probabilistically aggregating municipality-level predictions across 27 states. Municipality-level predictive distributions were aggregated using Monte Carlo sampling with a Gaussian copula to account for assumed dependence between municipalities (see Methods). Negative ΔWIS values favour fitting directly to aggregated state-level case series. Lines connect the 1-, 3-, 6- and 12-month forecast horizons.

**Table 1.** Epidemiological, climatic and demographic covariates used in the dengue forecasting models. The full covariate specification included all covariates shown. The temporal specifications describe explicitly constructed covariates rather than the full historical input available to each forecasting model. Models additionally use historical dengue case sequences or autoregressive structure, as detailed in Table 2. The minimal covariate specification evaluated in the covariate-selection analysis comprised log(1 + cases) at lag 1, the mean of minimum temperature over lags 1–2, and the mean of precipitation over lags 1–2.

| **Covariate** | **Temporal specification** | **Data source** |
|---|---|---|
| Minimum temperature | Monthly values at lags 0, 1 and 2 | WorldClim 2.1 (28); historical: CRU-TS 4.09 (29), 2.5 arc-min; projected: CMIP6 ACCESS-CM2 (30–31), SSP2-4.5, 2.5 arc-min |
| Maximum temperature | Monthly values at lags 0, 1 and 2 | WorldClim 2.1 (28); historical: CRU-TS 4.09 (29), 2.5 arc-min; projected: CMIP6 ACCESS-CM2 (30–31), SSP2-4.5, 2.5 arc-min |
| Precipitation | Monthly values at lags 0, 1 and 2 | WorldClim 2.1 (28); historical: CRU-TS 4.09 (29), 2.5 arc-min; projected: CMIP6 ACCESS-CM2 (30–31), SSP2-4.5, 2.5 arc-min |
| Oceanic Niño Index (ONI) | Monthly value and 6- and 12-month right-aligned moving averages | NOAA Oceanic Niño Index (32) |
| Standardized Precipitation Index (SPI-6) | 6-month SPI | Global Drought Observatory (GDO), Copernicus Emergency Management Service (33) |
| Population | Annual population estimate assigned to each month of the corresponding year | WorldPop (34) |
| Dengue cases | log(1 + cases), lag 1 | InfoDengue (26) |

**Table 2.**
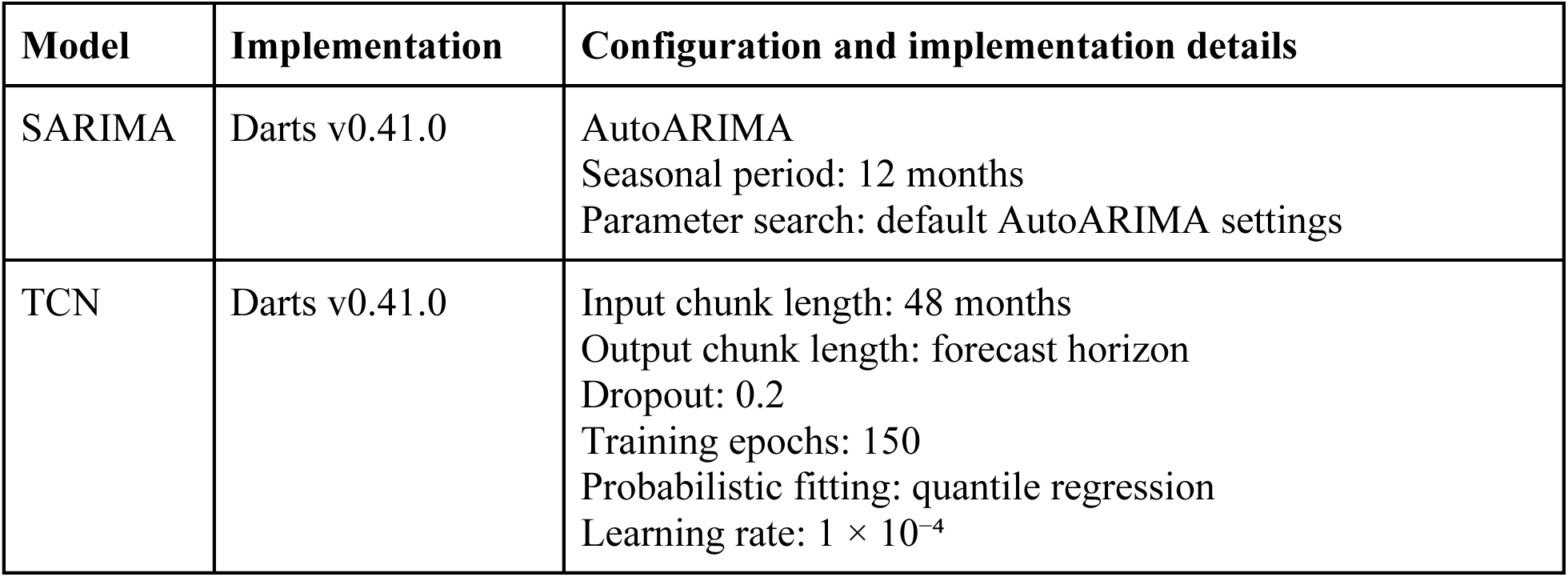
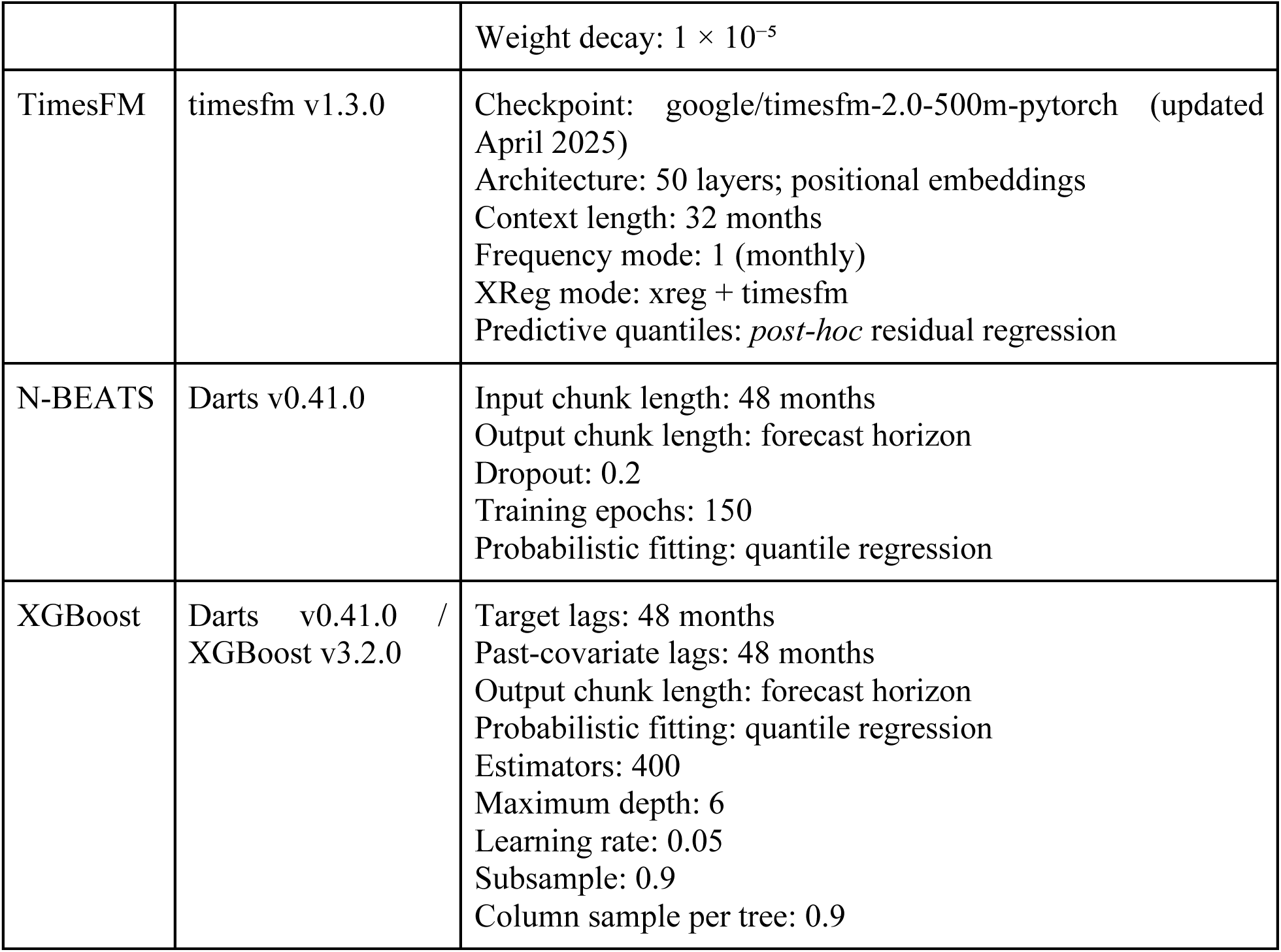
Forecasting models, implementations and specifications used in the primary analysis. Forecasts were generated and evaluated at 1-, 3-, 6- and 12-month horizons, using horizon-specific model configurations where applicable. Probabilistic forecasts used quantiles 0.01, 0.025, 0.05, 0.1, 0.2, 0.3, 0.4, 0.5, 0.6, 0.7, 0.8, 0.9, 0.95, 0.975 and 0.99. TimesFM was used without task-specific parameter fine-tuning, with predictive quantiles constructed using the post-hoc residual-regression procedure described below.

| Model | Implementation | Configuration and implementation details |
|---|---|---|
| SARIMA | Darts v0.41.0 | AutoARIMA<br>Seasonal period: 12 months<br>Parameter search: default AutoARIMA settings |
| TCN | Darts v0.41.0 | Input chunk length: 48 months<br>Output chunk length: forecast horizon<br>Dropout: 0.2<br>Training epochs: 150<br>Probabilistic fitting: quantile regression<br>Learning rate: 1 × 10<sup>−4</sup> |
|  |  | Weight decay: 1 × 10<sup>−5</sup> |
| TimesFM | timesfm v1.3.0 | Checkpoint: google/timesfm-2.0-500m-pytorch (updated April 2025)<br>Architecture: 50 layers; positional embeddings<br>Context length: 32 months<br>Frequency mode: 1 (monthly)<br>XReg mode: xreg + timesfm<br>Predictive quantiles: *post-hoc* residual regression |
| N-BEATS | Darts v0.41.0 | Input chunk length: 48 months<br>Output chunk length: forecast horizon<br>Dropout: 0.2<br>Training epochs: 150<br>Probabilistic fitting: quantile regression |
| XGBoost | Darts v0.41.0 / XGBoost v3.2.0 | Target lags: 48 months<br>Past-covariate lags: 48 months<br>Output chunk length: forecast horizon<br>Probabilistic fitting: quantile regression<br>Estimators: 400<br>Maximum depth: 6<br>Learning rate: 0.05<br>Subsample: 0.9<br>Column sample per tree: 0.9 |

To assess the effect of model fitting strategy, we compared municipality-level forecasts from models fitted independently to each municipality with forecasts for the same municipalities from models fitted jointly across municipalities within each state. The comparison included the 20 municipalities with the highest cumulative reported dengue case counts over the study period (Methods). The effect was strongly model-and horizon-dependent (Figure 3B; Supplementary Table S7). N-BEATS consistently and substantially favoured independent municipality-level fitting, with median municipality-level ΔWIS becoming more negative from −3.58 at one month to −10.57 at 12 months. TimesFM consistently favoured the state-grouped configuration, while TCN showed an advantage for state-grouped fitting at one month with little difference thereafter. XGBoost shifted in the opposite direction with horizon: effects were small at one and three months, while independent fitting was favoured at six (median ΔWIS = −0.78) and 12 months (−1.10). SARIMA showed little difference between fitting strategies.

Generating forecasts at coarser spatial scales introduces a distinct choice: whether to aggregate municipality-level case series before fitting a model, or to fit municipality-level models and subsequently aggregate their probabilistic forecasts. We compared these approaches across all 27 Brazilian states (Figure 3C; Supplementary Table S8). The effects of spatial aggregation were strongly model- and horizon-dependent. TCN consistently favoured aggregation of municipality-level forecasts, while TimesFM showed the same pattern with smaller effects. N-BEATS showed the opposite pattern: at one-month forecast horizon there was no consistent advantage for either aggregation strategy across states (median ΔWIS = −0.31; p = 0.25), whereas direct fitting to aggregated case series was favoured from three to 12 month horizons (median ΔWIS = −3.15 to −5.30; Supplementary Table S8). XGBoost shifted from favouring aggregation of municipality-level forecasts at one month horizons (median ΔWIS = +0.78) to favouring direct fitting to aggregated case series at six and 12 months horizons (−0.49 and −1.16, respectively). SARIMA differences were comparatively small and inconsistent across horizons.

### Monthly iterative retraining did not consistently improve forecast performance

Updating forecasting models as new observations become available may improve performance by incorporating recent epidemic dynamics, but frequent retraining incurs significant additional computational cost. We therefore compared the initial training regime, in which models were fitted once using data from January 2010 to December 2019, with monthly iterative retraining. For computational tractability, this secondary analysis was restricted to municipality-level experiments in Ceará and São Paulo. TimesFM was excluded because its pretrained model parameters were not fitted or fine-tuned to the study data, making iterative retraining inapplicable. SARIMA showed little difference between training strategies in either state, whereas XGBoost consistently favoured initial training across all horizons in both states (Supplementary Table S9). TCN similarly favoured initial training across all horizons in Ceará. A valid N-BEATS retraining comparison could not be made in either state due to numerical instability. Overall, monthly retraining provided no consistent performance advantage among the models and settings for which valid comparisons were available.

### AI-guided model development independently selects PDFM embeddings for dengue forecasting

In ERA model development, we used data available through August 2023 and optimised mean WIS across 1-, 3-, 6- and 12-month forecast horizons. The resulting ERA-derived forecasting model was then frozen and evaluated across all Brazilian municipalities during an independent hold-out period from September 2023 to August 2025 (Methods). The ERA-derived model did not consistently outperform the primary forecasting models across horizons (Supplementary Table S10). However, ERA independently selected temporal epidemic dynamics, spatial propagation, climatic information and PDFM embeddings in its final forecasting model (Supplementary Table S11). The selection of PDFM embeddings through this independent model-development process provides additional evidence that their predictive contribution is not specific to the residual-adjustment approach used in the primary analyses.

## Discussion

Our findings demonstrate how pretrained temporal and spatial representations can contribute to probabilistic infectious disease forecasting, while highlighting the importance of forecasting architecture and modelling configuration in determining their predictive value. Across Brazil, the relative performance of the five architectures evaluated depended strongly on forecast horizon, while the benefits of PDFM residual adjustment varied according to the underlying model and epidemiological setting. These findings extend previous dengue forecasting research demonstrating substantial variation in predictive performance across models, locations and epidemic seasons (12–15), and suggest that the value of pretrained representations depends not only on the information they encode but also on how that information is incorporated into forecasting systems.

The comparatively stable performance of TimesFM at longer horizons is particularly relevant given the challenges of forecasting dengue dynamics beyond the short term (12, 15). Whereas most architectures evaluated showed deteriorating probabilistic performance with increasing horizon, TimesFM maintained relatively stable WIS across 3-, 6- and 12-month forecasts. One possible explanation is that large-scale temporal pretraining enables the model to exploit patterns learned across diverse time series, potentially supporting generalisation beyond the historical dynamics represented in the Brazilian dengue training data. However, this mechanism cannot be established from the present comparisons, and the observed advantage may also reflect differences in model architecture, probabilistic forecast construction or the configurations evaluated. TimesFM’s performance should therefore be interpreted as evidence for the potential utility of temporal pretraining, rather than demonstrating its superiority over other dengue forecasting approaches.

The potential value of maintaining forecasting performance at seasonal to annual timescales is particularly apparent in the context of dengue’s geographical expansion. The record 2024 epidemics in Brazil and across the Americas, coinciding with El Niño-associated temperature anomalies and expansion into previously less-affected regions (4), highlight the challenges of anticipating changing epidemic dynamics. Reliable longer-horizon forecasts could support earlier public health preparedness, including advance allocation of resources and planning of vector-control and healthcare responses. However, their operational value will depend on performance under changing epidemiological conditions and the availability of timely surveillance data.

The model-dependent benefits of PDFM adjustment suggest that pretrained spatial representations can provide predictive information beyond historical dengue cases and conventional covariates, although their utility depends on the forecasting architecture and the errors remaining in its predictions. PDFM embeddings capture aspects of population activity, connectivity, the built environment and environmental context that may not be fully represented by conventional predictors. However, larger improvements following adjustment do not necessarily indicate that these representations contain more useful information for a particular model or setting. Our WIS decomposition showed that improvements arose primarily through reductions in out-of-interval penalties, suggesting that PDFM adjustment may be particularly valuable when baseline predictive distributions fail to capture observed epidemic outcomes. The geographical analyses further demonstrated that regional differences in absolute WIS improvements were sometimes attenuated when evaluated using log-relative skill, consistent with a contribution from the magnitude and distribution of baseline forecast errors to the apparent heterogeneity. This interpretation is consistent with the greater improvements observed at higher reported dengue case counts, where forecast errors may also be larger. Together, these findings suggest that PDFM adjustment benefits depend on the interaction between contextual information, forecasting architecture and residual-error structure, although the specific mechanisms and embedding components responsible remain unresolved.

Geographical variation in baseline forecasting performance was also evident, particularly in southern Brazil. Although TCN achieved the lowest mean one-month WIS across all five macroregions, its advantage over TimesFM was substantially weaker in Rio Grande do Sul and Santa Catarina than elsewhere. This raises the possibility that pretrained temporal representations may be comparatively useful where epidemic dynamics differ from historical patterns, although our exploratory analyses of historical dengue activity and changes in reported cases and population-adjusted incidence did not establish a consistent association with relative model performance. Geographical differences were also apparent in PDFM adjustment benefits, particularly for N-BEATS and XGBoost, suggesting that the contribution of residual adjustment varies across epidemiological settings as well as forecasting architectures. Together, these findings reinforce the importance of evaluating forecasting approaches across diverse geographical and epidemiological conditions (15).

The model-dependent effects of covariate selection and spatial fitting suggest that forecasting architecture and modelling configuration should be considered jointly rather than as independent choices. Additional covariates may improve performance when an architecture can exploit their predictive information, but may provide little benefit or introduce unnecessary complexity when historical epidemic dynamics already capture much of the relevant variation. Similarly, joint fitting across municipalities may allow models to exploit shared temporal patterns, whereas independent fitting may better preserve locally specific dynamics. Our results indicate that neither approach is consistently preferable across the architectures evaluated, highlighting the importance of assessing configuration choices within individual forecasting systems rather than assuming that greater model complexity or information sharing necessarily improves performance. The comparison of state-level forecast aggregation strategies further demonstrates that the preferred approach depends on the forecasting architecture and horizon.

Together, these findings highlight substantial variation in forecasting performance across the models, horizons and modelling configurations evaluated, arguing against a single optimal strategy. Our comparisons contribute to existing evidence on dengue forecasting in Brazil by informing choices concerning forecasting architecture, horizon, covariate selection, spatial fitting and forecast aggregation (Supplementary Table S12). The observed heterogeneity motivates exploring ensemble approaches that combine forecasts from different models or configurations, potentially exploiting complementary strengths across epidemiological settings and forecast horizons. Evaluating whether such combinations improve predictive performance, and how their contributions should be weighted across locations and horizons, represents an important direction for future research.

The absence of consistent benefits from monthly retraining highlights a potential trade-off between incorporating recent epidemiological information and maintaining stable forecasting performance. Although frequent updating may allow models to respond to changing epidemic dynamics, it may also introduce variability in fitted parameters without improving predictive accuracy, particularly when recent observations reflect short-term fluctuations rather than persistent changes in transmission. The numerical instability encountered with N-BEATS further illustrates that computational and implementation considerations can influence the feasibility of frequent model updating. These findings suggest that retraining frequency should be evaluated as a model-specific design choice, balancing potential improvements in responsiveness against forecast accuracy, computational cost and numerical stability. However, because our experiments compared only initial training with monthly retraining in Ceará and São Paulo, they do not establish whether alternative updating schedules or adaptation strategies would improve performance.

The ERA experiment provides an independent perspective on the predictive value of PDFM embeddings. Unlike the primary analyses, which incorporated PDFM through post-hoc residual adjustment, ERA independently selected embedding dimensions to parameterise municipality-specific characteristics of its forecasting model, including climate sensitivities and forecast uncertainty. This suggests that PDFM-derived information may contribute to dengue forecasting through modelling approaches beyond residual correction, although neither approach identifies which components of the multimodal representation are responsible for the predictive signal. The ERA-derived model did not consistently outperform the primary architectures over the hold-out period, but its independent selection of PDFM embeddings provides further support for their contribution to dengue forecasting.

More broadly, our findings illustrate three distinct roles for AI in infectious-disease forecasting. Time-series foundation models such as TimesFM can transfer representations learned through large-scale temporal pretraining, potentially supporting forecasting across longer horizons. Geospatial foundation models such as PDFM can provide representations of human and environmental context that augment disease-specific forecasting models, although their benefits depend on the underlying architecture and how these representations are incorporated. AI-guided model development offers a third approach, identifying and combining predictive information within models developed for a particular task. These approaches are not necessarily interchangeable, and their relative value will depend on the forecasting objective, available information and modelling framework. Together, they demonstrate different ways in which AI can contribute to epidemic forecasting, while highlighting the need to evaluate their benefits within specific epidemiological and operational settings.

The variation in forecasting performance across architectures, horizons and modelling configurations highlights the need for flexible, reproducible frameworks that support systematic comparison of forecasting approaches. The modular Thucia framework (Methods) used in this study was designed to facilitate such comparisons by separating data acquisition, model fitting, probabilistic forecasting, residual adjustment and evaluation into interchangeable components. This flexibility is particularly relevant when forecasting objectives, data availability and operational requirements differ across geographical settings or change over time. More broadly, our findings reinforce the importance of benchmarks that evaluate forecasting approaches across different epidemiological contexts, spatial scales and modelling configurations, rather than relying on performance under a single experimental setting. Such evaluations could help inform model selection and adaptation for operational forecasting, while establishing whether the context-dependent patterns observed in Brazil generalise to other pathogens, geographical settings and surveillance systems.

### Limitations and future directions

We acknowledge several limitations. First, our primary analysis is restricted to Brazil; although this provides a stringent test across diverse epidemiological and climatic settings, generalisability to other countries, pathogens and surveillance systems requires further evaluation. Second, although we examined correlations among PDFM embedding dimensions and their associations with external spatial characteristics, we did not isolate the contributions of individual embedding components to forecasting performance. The latent nature of these representations limits mechanistic interpretation, and further component-specific analyses would be needed to establish which behavioural, built-environment or environmental characteristics drive the observed improvements. Third, the forecasting architectures were selected to represent distinct modelling paradigms rather than to provide an exhaustive benchmark of dengue forecasting methods. Modelling configurations and hyperparameters were selected pragmatically rather than through exhaustive optimisation, and alternative choices could alter the relative performance of individual models. Comparisons with additional dengue-specific statistical models and established ensemble approaches would help establish whether the benefits of pretrained representations generalise to other forecasting systems. The comparison of independent municipality-level and joint state-grouped fitting was also restricted to 20 municipalities with high cumulative reported dengue case counts, limiting the extent to which these configuration-specific findings can be generalised across Brazil. Direct comparison with recent initiatives such as IMDC24 is further complicated by differences in surveillance outcomes, spatial and temporal resolution, forecast origins and evaluation protocols. In particular, IMDC24 evaluated weekly forecasts of probable dengue cases at state level over epidemiological seasons, whereas our study evaluates monthly forecasts of notified cases at municipality level across fixed forecast horizons. A further limitation concerns the potential overlap between TimesFM’s pretraining data and the data used in this study. Although TimesFM was applied without task-specific parameter fine-tuning, potential overlap between the study data and its pretraining corpus cannot be completely ruled out because the composition of the pretraining data is not exhaustively identifiable at the individual-series level. Fourth, the retraining analysis was restricted to monthly updating in Ceará and São Paulo, and a valid comparison could not be made for N-BEATS because of numerical instability. Finally, all primary analyses were retrospective and depended on the quality and completeness of the underlying surveillance data. Historical reporting vintages were not reconstructed, and forecasts were generated and evaluated using a frozen retrospective InfoDengue extraction in which historical case counts may incorporate revisions made after the corresponding forecast origin. The evaluation also encompassed the COVID-19 pandemic and subsequent large dengue epidemics, during which epidemiological and surveillance changes may have affected relationships learned from historical data. Furthermore, historical PDFM embeddings aligned to individual forecast origins were unavailable, and a single October 2023 snapshot was used throughout the evaluation period. These limitations mean that the findings should be interpreted as retrospective evidence rather than established operational forecasting performance.

### Conclusion

In conclusion, our findings demonstrate the potential of pretrained temporal and spatial representations to strengthen probabilistic dengue forecasting in Brazil. Among the five architectures evaluated, TimesFM maintained stronger relative performance at longer forecast horizons, while PDFM-derived spatial representations provided additional predictive information for several models through residual adjustment. However, these benefits depended on forecasting architecture, horizon and geographical setting, with modelling configuration further influencing predictive performance. Together, these findings highlight the importance of evaluating forecasting approaches across different epidemiological and operational contexts, rather than assuming that any single architecture or configuration will perform consistently best. Integrating pretrained representations into dengue forecasting systems therefore offers opportunities to improve predictive performance, but their practical value will require further validation under operational forecasting conditions.

## Materials & Methods

### Epidemiological data

Municipality-level dengue case data were obtained from the InfoDengue application programming interface (API) (26) for January 2010 to August 2025. Data were extracted on 10 November 2025 using Brazilian Institute of Geography and Statistics (IBGE) municipality codes. We used notified dengue cases (*casos*), rather than probable dengue cases or InfoDengue nowcast estimates. Weekly reported cases were assigned to the calendar month containing the start of the corresponding epidemiological week and summed to obtain monthly reported case counts, with monthly totals comprising four or five epidemiological weeks. This aggregation was applied consistently across all forecasting models. Municipalities were subsequently aligned to Global Administrative Areas Database (GADM; 27) administrative level-2 regions, and the resulting dataset was frozen for all analyses. Historical reporting vintages were not reconstructed, so case counts reflect the values available at extraction rather than necessarily those available at each forecast origin.

### Forecast training and evaluation

SARIMA, TCN, XGBoost and N-BEATS were fitted once using data from January 2010 through December 2019, after which the fitted models were held fixed while generating forecasts at 1-, 3-, 6- and 12-month horizons. Before model fitting, spatial units with no reported dengue cases during the January 2010–December 2019 training period were excluded from the fitting dataset. Forecast evaluation was defined by target month, with a common evaluation window of January 2020– August 2025 at each horizon. Consequently, for early target months at longer horizons, the nominal forecast origin precedes the December 2019 model-fitting cutoff; these forecasts should therefore be interpreted as horizon-specific retrospective predictions from models fitted to a fixed training period, rather than strictly prospective forecasts using only information available at the nominal forecast origin. TimesFM was applied without fitting or fine-tuning its pretrained model parameters to the study data. Forecasts targeting months after August 2025 were excluded from evaluation because corresponding observed outcomes were unavailable.

### Covariate data

Epidemiological, climatic and demographic covariates used in the forecasting models are summarised in Table 1. For consistency across forecasting models, all covariate time series were treated as past covariates, with projected values substituted where required.

### Modelling framework

We conducted all analyses using Thucia (https://github.com/kraemer-lab/Thucia), a modular open-source forecasting framework developed to standardise the acquisition, aggregation, modelling and evaluation of epidemiological data across spatial and temporal scales. Thucia separates data engineering, model fitting, probabilistic forecasting, residual adjustment and evaluation into interchangeable components, allowing different models and modelling strategies to be evaluated within a consistent experimental workflow. In the present analysis, Thucia was used to implement the five forecasting models and the different covariate, spatial fitting, aggregation and retraining configurations described below.

We selected five forecasting models spanning distinct statistical, machine-learning, deep-learning and foundation-model paradigms. SARIMA provides a conventional statistical approach to modelling autoregressive and seasonal dynamics. TCN and N-BEATS provide complementary deep-learning architectures for modelling temporal dependencies (35, 36), while XGBoost represents flexible nonlinear machine learning using lagged reported dengue case counts and environmental covariates (37). TimesFM allows us to evaluate the potential benefits of large-scale time-series pretraining (21). This selection was intended to assess whether the predictive contribution of PDFM embeddings generalises across different forecasting architectures, rather than to exhaustively benchmark all available dengue forecasting methods. We evaluated forecasts at monthly resolution to assess predictive performance across seasonal to annual horizons relevant to public health preparedness, while maintaining a consistent temporal resolution across forecasting architectures. Forecasts were evaluated at 1-, 3-, 6- and 12-month horizons (see Forecast training and evaluation). Where supported by the model, multi-step forecasts were generated jointly over the corresponding horizon rather than by recursively feeding one-month-ahead predictions back into the model. Forecast horizons denote complete calendar-month lead times between the nominal forecast origin and target month. For example, a July 2024 target evaluated at the three-month horizon represents incident cases during July 2024 at a nominal three-month forecast lead.

In the primary analysis, municipality-level time series were grouped by state. TCN, N-BEATS and XGBoost were fitted jointly across municipalities within each state, allowing model parameters to be shared across local time series. SARIMA was fitted independently to each municipality, reflecting its implementation as a univariate statistical model, while TimesFM generated forecasts from individual municipality-level histories using pretrained model parameters without task-specific fine-tuning. These approaches reflect differences in how the forecasting architectures accommodate multiple time series, rather than an attempt to optimise the spatial fitting strategy separately for each model.

Model hyperparameter tuning was intentionally limited to evaluate established forecasting architectures under computationally feasible configurations, rather than optimising each model exhaustively for the study setting. Where choices were required, study-specified settings were selected pragmatically based on established time-series modelling practice, computational tractability and exploratory runs on training-period data using the 645 municipalities of São Paulo state. For example, neural and tree-based models used 48 months of historical input, while SARIMA used a 12-month seasonal period; SARIMA model orders were otherwise selected automatically using AutoARIMA. These settings were subsequently held fixed in the primary Brazil-wide analyses rather than optimised separately by municipality or against the evaluation period. Several broader modelling choices, including covariate specification, independent municipality-level versus joint state-grouped fitting, spatial aggregation and retraining, were evaluated explicitly as configuration analyses rather than selected through hyperparameter optimisation. Table 2 reports both study-specified settings and fixed or automatically determined properties of the model implementations.

Covariate time series were scaled by their mean absolute value calculated over the training period to maintain comparable magnitudes and improve numerical stability. For the primary forecasting analyses, case counts were transformed as log(1 + cases) for model fitting and WIS evaluation. SARIMA, TCN, N-BEATS and XGBoost were implemented using the Darts time-series forecasting library (38), with XGBoost accessed through the corresponding Darts forecasting interface.

### PDFM embeddings and residual adjustment

We assessed whether Population Dynamics Foundation Model (PDFM) embeddings provided predictive information complementary to the baseline forecasting models (17). Historical PDFM embeddings aligned to individual forecast origins were not available; therefore, a single October 2023 embedding snapshot was used throughout the evaluation period. PDFM embeddings were available for 5,568 of the 5,570 municipalities included in the baseline analysis. Passabém (Minas Gerais) and Pindoba (Alagoas), were absent from the provided embeddings and so were excluded from PDFM-adjusted comparisons.

To apply PDFM consistently across forecasting models, we used a residual-regression adjustment on the log(1 + cases) scale. For each forecast horizon and predictive quantile q, residuals were defined as the observed log-transformed case count minus the corresponding forecast quantile. A separate histogram-based gradient-boosting regression model with quantile (pinball) loss was fitted for each predictive quantile using municipality-level PDFM embeddings as predictors. For each target month, residual models were fitted using observations from a 24-month sliding window ending at the corresponding nominal forecast origin (target month minus forecast horizon), such that outcome data after the nominal forecast origin were excluded. The predicted residual was added to the corresponding forecast quantile, after which adjusted forecasts were transformed back to the original case-count scale. PDFM embeddings themselves were static and were not temporally aligned to individual forecast origins.

To illustrate the structure of the PDFM representation (Figure 1C–D), pairwise Pearson correlations were calculated across the 330 embedding dimensions. The representation comprises three modality-specific components: aggregated Search Trends (dimensions 0–127), Maps & Busyness (dimensions 128–255), and Weather & Air Quality (dimensions 256–329). For visualisation, one representative dimension from each component was selected as that with the largest absolute municipality-level Pearson correlation with a prespecified external spatial characteristic: log-transformed nighttime light intensity (39), population and temperature, respectively. These correlations were used only to illustrate embedding structure and were not used in forecast-model fitting or evaluation.

### Forecast modelling configuration analyses

We conducted three analyses to assess how modelling configuration influenced forecast performance. First, we compared the full covariate specification with the minimal covariate set comprising log(1 + cases) at lag 1, the mean of minimum temperature over lags 1–2, and the mean of precipitation over lags 1–2 (Table 1). Second, for the 20 municipalities with the highest cumulative reported dengue case counts over the study period, we compared municipality-level forecasts from models fitted independently to each municipality with forecasts for the same municipalities from models fitted jointly across municipalities within each state. Third, we compared two approaches to generating state-level forecasts across all 27 Brazilian states: fitting models directly to state-level aggregated case series and aggregating municipality-level probabilistic forecasts after model fitting.

For the state-level forecast-aggregation analysis, municipality-level predictive distributions were combined using Monte Carlo simulation because quantiles of municipality-level forecasts cannot be directly summed to obtain quantiles of the state-level predictive distribution. For each municipality, a predictive distribution was approximated by interpolation between forecast quantiles. Joint draws from municipality-level distributions were generated using a Gaussian copula with a fixed dependence parameter of ρ = 0.3, selected pragmatically to represent moderate positive dependence between municipality-level forecasts without assuming either independence or strong spatial dependence. Simulated municipality-level case counts were summed within each state, and quantiles of the resulting simulated state-level distribution were used as the aggregated probabilistic forecast. Observed state-level case counts were obtained by summing reported municipality-level cases.

For each of the three modelling-configuration comparisons, forecasts were paired by spatial unit, target month and forecast horizon. Differences in WIS were calculated between paired configurations and averaged across the January 2020–August 2025 target-month evaluation period within each spatial unit. The resulting municipality-or state-level mean differences were summarised by their mean, median and interquartile range. Two-sided Wilcoxon signed-rank tests were applied to the paired spatial-unit summaries where inferential comparisons were reported. Thus, the inferential unit for these analyses was the municipality or state after averaging forecast-performance differences through time.

As a separate secondary analysis, we evaluated monthly iterative retraining in municipality-level experiments in Ceará and São Paulo. Under the initial training regime, each model was fitted once using data from January 2010 to December 2019 and was not subsequently retrained during the evaluation period. Under monthly retraining, models were refitted at each monthly forecast interval using an expanding training window, retaining the original January 2010 start date and incorporating all observations available up to the corresponding forecast origin. WIS was averaged across the January 2020–August 2025 target-month evaluation period within each municipality, and paired municipality-level means were compared using two-sided Wilcoxon signed-rank tests. Retraining comparisons were reported only for model–state combinations for which usable paired forecasts were available; numerical failures were recorded separately. TimesFM was excluded because its pretrained model parameters were not fitted or fine-tuned to the study data, making iterative retraining inapplicable.

### Model evaluations

Models were evaluated primarily using the Weighted Interval Score (WIS), a proper scoring rule for probabilistic forecasts (40) that assesses predictive calibration and sharpness. WIS was calculated on the log(1 + cases) scale to reduce the influence of differences in absolute reported case counts across municipalities and time. Lower WIS values indicate better probabilistic forecast performance. The coefficient of determination (R²) was used as a complementary measure of agreement between point predictions and observed reported case counts.

For the analysis in Figure 2C, we constructed a seasonal reference forecast separately for each municipality and calendar month using only observations from the January 2010–December 2019 training period. For each municipality–calendar-month combination, the seasonal reference forecast was defined as the mean log(1 + cases) across the corresponding calendar month in the training years. This value was used for all predictive quantiles, yielding a degenerate predictive distribution centred on the historical seasonal mean, and was held fixed throughout the January 2020–August 2025 evaluation period. The baseline was scored using the same quantile grid and WIS implementation as the forecasting models. For each matched municipality and target month, model skill relative to the seasonal baseline was calculated as log(1 + WIS_seasonal_) − log(1 + WIS_model_), such that positive values indicate lower WIS, and therefore better probabilistic forecast performance, for the forecasting model.

To characterise how PDFM adjustment affected probabilistic forecast performance, we separated the constituent WIS terms (40) into contributions from median forecast error, predictive-interval width and out-of-interval penalties. For each paired PDFM-adjusted and base forecast, the change in each component was calculated as the PDFM-adjusted contribution minus the corresponding base-forecast contribution, such that negative values indicate an improvement. Component-wise differences were then averaged across municipality-month forecasts; their sum equals the total mean ΔWIS.

To examine whether the effect of PDFM adjustment varied with observed dengue case counts, municipality-month forecasts were stratified retrospectively according to the number of reported cases in the forecast target month. Observations with zero reported cases formed a separate stratum. Within each target month and model–horizon combination, observations with positive reported case counts were divided into low, moderate and high strata using the 33rd and 67th percentiles of their case-count distribution. For each stratum, we calculated the proportion of paired forecasts for which PDFM adjustment reduced WIS (ΔWIS < 0). To quantify the magnitude of these effects, mean ΔWIS was calculated within each stratum and target month, then averaged equally across target months. High-minus-low contrasts were calculated as paired differences between monthly mean ΔWIS values in the highest and lowest positive case-count strata. Uncertainty was estimated using a temporal moving-block bootstrap with three-month blocks and 2,000 replicates, with sensitivity analyses using six- and twelve-month blocks. These stratified analyses were exploratory, and confidence intervals were not adjusted for multiple comparisons.

For comparisons involving PDFM residual adjustment, mean differences in WIS were evaluated using a temporal moving-block bootstrap (41) to account for chronological dependence. For each model and forecast horizon, ΔWIS was first averaged across municipalities within each target month to form a chronological sequence, from which contiguous blocks of three months were resampled with replacement. The 95% confidence interval was defined by the 2.5th and 97.5th percentiles of 2,000 bootstrap replicates. Sensitivity analyses used block lengths of 6 and 12 months. As a complementary test of predictive accuracy, we applied two-sided Diebold–Mariano tests (42, 43) to chronological target-month sequences of mean WIS, obtained by averaging WIS across municipalities within each target month. The dependence parameter was set to the forecast horizon, with a Bartlett variance estimator and Harvey small-sample correction. To account for multiple testing across the 20 model–horizon comparisons, statistical significance was assessed using a Bonferroni-adjusted threshold of α = 0.0025 (0.05/20).

SARIMA produced occasional extreme predictions in a small number of municipalities, particularly at longer forecast horizons. Across the evaluation period, the forecast median exceeded one million cases in 13 of 1,515,584 municipality-month-horizon forecasts (<0.001%): none at the 1-month horizon, one at 3 months, three at 6 months, and nine at 12 months. These finite extreme forecasts were retained in the quantitative WIS evaluation to avoid artificially improving estimated forecast performance, but were excluded from selected visualisations where their magnitude would otherwise obscure the displayed forecast trajectories.

### AI-guided model development

As a complementary analysis, we used Google’s Empirical Research Assistance (ERA) to develop a probabilistic dengue forecasting model across all Brazilian municipalities. ERA was provided with historical reported dengue case counts, climatic and ecological covariates, municipality-level spatial adjacency and static PDFM embeddings, and was tasked with minimising mean Weighted Interval Score (WIS) across 1-, 3-, 6-and 12-month forecast horizons. Model development preserved temporal ordering of the epidemiological observations, such that forecasts at each origin used only observations assigned to that date or earlier; the static PDFM embeddings were fixed across forecast origins as described above. Model development and optimisation used target observations through August 2023. The resulting forecasting function was then frozen and evaluated against the September 2023–August 2025 hold-out period, which was not used during model development or optimisation.

The final ERA-derived model used municipality- and month-specific historical reported case counts to define seasonal baselines, adjusted according to recent deviations from local seasonal patterns. Recent epidemic momentum was partially propagated between neighbouring municipalities through the spatial adjacency network and progressively damped with increasing forecast horizon. Climatic information was incorporated through the Oceanic Niño Index (ONI) and six-month Standardized Precipitation Index (SPI-6). Selected dimensions of the static PDFM embeddings parameterised municipality-specific climate sensitivities, baseline dispersion and forecast volatility (Supplementary Table S11). Expected case counts were converted to probabilistic forecasts using Negative Binomial distributions, with dispersion varying by municipality and forecast horizon. Quantile forecasts were generated at each of the four horizons and evaluated using WIS over the independent hold-out period.

## Data Availability

Dengue surveillance data were obtained from the publicly accessible InfoDengue API (https://info.dengue.mat.br/). Climatic, demographic and environmental data were obtained from the publicly available sources described in the manuscript. Processed data and scripts used to generate the figures and tables are available at https://github.com/kraemer-lab/pdfm-dengue-brazil. The forecasting framework is available at https://github.com/kraemer-lab/Thucia. Population Dynamics Foundation Model (PDFM) embeddings are available for research use subject to approval by Google Research (https://github.com/google-research/population-dynamics). Access to the embeddings and other third-party datasets is subject to the respective providers' terms and conditions.

## Acknowledgements

We thank Cláudia Torres Codeço for feedback on this manuscript. M.U.G.K. acknowledges funding from The Rockefeller Foundation (PC-2022-POP-005), Health AI Programme from Google.org, the Oxford Martin School Programmes in Pandemic Genomics & Digital Pandemic Preparedness, European Union’s Horizon Europe programme projects MOOD (#874850) and E4Warning (#101086640), Wellcome Trust grants 303666/Z/23/Z, 226052/Z/22/Z & 228186/Z/23/Z, the United Kingdom Research and Innovation (#APP8583), the Medical Research Foundation (MRF-RG-ICCH-2022-100069), UK International Development (301542–403), the Bill & Melinda Gates Foundation grants (INV-063472, INV-090281, INV-103122) and Novo Nordisk Foundation (NNF24OC0094346). The contents of this publication are the sole responsibility of the authors and do not necessarily reflect the views of the European Commission or the other funders.

## Code and data availability

The forecasting analyses were conducted using the open-source Thucia forecasting framework, available at https://github.com/kraemer-lab/Thucia. Processed data and scripts used to generate the figures and tables presented in this study are publicly available at https://github.com/kraemer-lab/pdfm-dengue-brazil. Dengue surveillance data were obtained through InfoDengue (26), with extraction and processing described in Methods; sources of climatic, demographic and environmental covariates are described in Table 1. PDFM embeddings and other data used in the study are available subject to the access and redistribution conditions of their respective providers.

