## Supplementary for "Foundation models capture human and environmental context to improve dengue forecasting"

### Supplementary materials

**Table S1. Baseline forecasting performance across models and forecast horizons.** SARIMA, TCN, XGBoost and N-BEATS were fitted using data from January 2010 to December 2019; TimesFM was applied without task-specific parameter fine-tuning. All models were evaluated retrospectively over a common January 2020–August 2025 target-month evaluation period spanning 5,570 Brazilian municipalities. WIS and $R^2$ are summarised across municipality-month forecasts (n = 378,760 per model–horizon combination). Lower WIS indicates better probabilistic forecast performance; higher $R^2$ indicates greater correspondence between predicted and observed case counts. Mean, median and 95th-percentile WIS are reported to summarise both typical performance and the upper tail of forecast errors.

| Model | Horizon (months) | Mean WIS | Median WIS | P95 WIS | Mean $R^2$ |
|---|---|---|---|---|---|
| SARIMA | 1 | 1.455 | 0.864 | 5.124 | 0.469 |
| SARIMA | 3 | 2.304 | 1.264 | 8.889 | 0.061 |
| SARIMA | 6 | 2.548 | 1.393 | 9.762 | 0.023 |
| SARIMA | 12 | 2.491 | 1.401 | 9.332 | 0.022 |
| TCN | 1 | 1.273 | 0.888 | 4.191 | 0.521 |
| TCN | 3 | 1.885 | 1.177 | 6.663 | 0.177 |
| TCN | 6 | 2.082 | 1.256 | 7.525 | 0.128 |
| TCN | 12 | 2.157 | 1.288 | 7.778 | 0.115 |
| XGBoost | 1 | 1.376 | 0.848 | 4.825 | 0.534 |
| XGBoost | 3 | 2.029 | 1.054 | 8.154 | 0.166 |
| XGBoost | 6 | 2.281 | 1.111 | 9.382 | 0.073 |
| XGBoost | 12 | 2.409 | 1.126 | 9.956 | 0.051 |
| N-BEATS | 1 | 2.146 | 0.909 | 8.478 | 0.249 |
| N-BEATS | 3 | 3.028 | 1.014 | 12.651 | 0.046 |
| N-BEATS | 6 | 3.259 | 1.036 | 13.555 | 0.033 |
| N-BEATS | 12 | 3.528 | 1.066 | 14.557 | 0.026 |

| | | | | | |
|---|---|---|---|---|---|
| TimesFM | 1 | 1.457 | 0.892 | 5.146 | 0.445 |
| TimesFM | 3 | 1.558 | 0.927 | 5.721 | 0.328 |
| TimesFM | 6 | 1.528 | 0.927 | 5.42 | 0.364 |
| TimesFM | 12 | 1.508 | 0.939 | 5.292 | 0.357 |

**Table S2. Geographical variation in baseline probabilistic dengue forecasting performance across Brazil.** Mean Weighted Interval Score (WIS) for SARIMA, TCN, XGBoost, N-BEATS and TimesFM at 1-, 3-, 6- and 12-month forecast horizons across Brazil's five macroregions. Additional comparisons between TCN and TimesFM are presented for Paraná, Rio Grande do Sul and Santa Catarina to characterise geographical variation within southern Brazil. Lower WIS indicates better probabilistic forecasting performance. Asterisks identify the lowest mean WIS at each horizon among the models included in the corresponding geographical comparison. Results cover 5,570 municipalities evaluated between January 2020 and August 2025.

| Geography | Model | Municipalities | 1 month WIS | 3 month WIS | 6 month WIS | 12 month WIS |
|---|---|---|---|---|---|---|
| North | SARIMA | 450 | 1.356 | 1.941 | 2.134 | 2.148 |
| | TCN | | 1.249* | 1.713 | 1.843 | 1.878 |
| | XGBoost | | 1.379 | 1.915 | 2.156 | 2.292 |
| | N-BEATS | | 2.015 | 2.61 | 2.785 | 2.965 |
| | TimesFM | | 1.44 | 1.465* | 1.455* | 1.470* |
| Northeast | SARIMA | 1794 | 1.305 | 1.878 | 2.035 | 2.022 |
| | TCN | | 1.134* | 1.581 | 1.724 | 1.769 |
| | XGBoost | | 1.208 | 1.667 | 1.83 | 1.909 |
| | N-BEATS | | 1.761 | 2.393 | 2.546 | 2.722 |
| | TimesFM | | 1.356 | 1.362* | 1.357* | 1.367* |
| Central-West | SARIMA | 467 | 1.645 | 2.526 | 2.807 | 2.727 |

| | | | | | | |
|---|---|---|---|---|---|---|
| | TCN | | 1.519* | 2.256 | 2.54 | 2.641 |
| | XGBoost | | 1.612 | 2.376 | 2.721 | 2.885 |
| | N-BEATS | | 2.179 | 3.18 | 3.486 | 3.717 |
| | TimesFM | | 1.722 | 1.799* | 1.771* | 1.769* |
| Southeast | SARIMA | 1668 | 1.627 | 2.794 | 3.169 | 3.101 |
| | TCN | | 1.383* | 2.156 | 2.429 | 2.507 |
| | XGBoost | | 1.488 | 2.337 | 2.677 | 2.815 |
| | N-BEATS | | 2.46 | 3.737 | 4.092 | 4.489 |
| | TimesFM | | 1.596 | 1.773* | 1.729* | 1.697* |
| South | SARIMA | 1191 | 1.404 | 2.311 | 2.504 | 2.378 |
| | TCN | | 1.240* | 1.885 | 2.048 | 2.165 |
| | XGBoost | | 1.379 | 2.051 | 2.28 | 2.451 |
| | N-BEATS | | 2.322 | 3.089 | 3.258 | 3.534 |
| | TimesFM | | 1.318 | 1.492* | 1.435* | 1.370* |
| Paraná | TCN | 399 | 1.518* | 2.401 | 2.707 | 2.856 |
| | TimesFM | | 1.75 | 1.954* | 1.885* | 1.821* |
| Rio Grande do Sul | TCN | 497 | 1.053* | 1.559 | 1.645 | 1.735 |
| | TimesFM | | 1.056 | 1.206* | 1.158* | 1.089* |
| Santa Catarina | TCN | 295 | 1.178 | 1.735 | 1.836 | 1.953 |
| | TimesFM | | 1.173* | 1.350* | 1.292* | 1.235* |

**Table S3. Heterogeneous effects of Population Dynamics Foundation Model (PDFM) adjustment on probabilistic dengue forecast performance.** Values are differences between PDFM-adjusted and unadjusted forecasts (ΔWIS = $WIS_{PDFM}$ − $WIS_{base}$, where base denotes the corresponding unadjusted forecast), such that negative values indicate improved probabilistic forecast performance. Mean ΔWIS and 95% confidence intervals were estimated using a temporal moving-block bootstrap with 3-month blocks and 2,000 bootstrap replicates, resampling contiguous blocks of chronological target-month mean ΔWIS. Sensitivity analyses using 6- and 12-month blocks produced generally consistent conclusions, with inference sensitive to block length only for TCN at 12 months and TimesFM at 3 months. The median municipality-level ΔWIS was calculated after averaging ΔWIS across the evaluation period within each municipality. "Municipalities improved" is the percentage of municipalities with a negative mean ΔWIS. Diebold–Mariano (DM) tests were applied to chronological target-month sequences of mean WIS, obtained by averaging across municipalities within each target month, using a two-sided horizon-specific lag structure, Bartlett variance estimator and Harvey small-sample correction. Positive DM statistics indicate lower WIS for PDFM-adjusted forecasts. The analysis covers the January 2020–August 2025 target-month evaluation period and 5,568 municipalities with available PDFM embeddings. At the 12-month TimesFM horizon, PDFM adjustment failed for 198 municipality-month forecasts (0.052% of potential paired observations); these observations were excluded from both the adjusted and corresponding unadjusted forecasts in paired comparisons. Diebold–Mariano p-values are reported without adjustment; statistical significance was assessed using a Bonferroni-corrected threshold of $\alpha = 0.0025$ to account for the 20 model–horizon comparisons. Bootstrap confidence intervals are unadjusted for multiple comparisons.

| Model | Horizon (months) | Mean ΔWIS (PDFM − base) | 95% temporal bootstrap CI | Median municipality-level ΔWIS | Municipalities improved (%) | DM statistic | DM p-value |
|---|---|---|---|---|---|---|---|
| N-BEATS | 1 | -0.5785 | [-0.7676, -0.4279] | -0.2524 | 91.6 | 10.082 | <0.0001 |
| N-BEATS | 3 | -1.0068 | [-1.3339, -0.7319] | -0.4655 | 89.9 | 6.484 | <0.0001 |
| N-BEATS | 6 | -1.1285 | [-1.4777, -0.8355] | -0.5027 | 89.5 | 5.65 | <0.0001 |
| N-BEATS | 12 | -1.1269 | [-1.5486, -0.7920] | -0.4876 | 88.5 | 3.389 | 0.001178 |
| SARIMA | 1 | -0.0556 | [-0.1028, -0.0241] | -0.0403 | 76.5 | 4.05 | 0.0001354 |
| SARIMA | 3 | -0.0776 | [-0.2039, 0.0112] | -0.0578 | 71.3 | 1.351 | 0.1812 |
| SARIMA | 6 | -0.0724 | [-0.2214, 0.0261] | -0.0588 | 67.4 | 0.926 | 0.3577 |
| SARIMA | 12 | -0.0121 | [-0.1207, 0.0583] | -0.0242 | 56.7 | 0.184 | 0.8545 |

| | | | | | | | |
|---|---|---|---|---|---|---|---|
| TCN | 1 | -0.0294 | [-0.0530, -0.0111] | -0.0046 | 62.6 | 3.803 | 0.0003113 |
| TCN | 3 | -0.081 | [-0.1487, -0.0263] | -0.0102 | 56.6 | 2.566 | 0.01253 |
| TCN | 6 | -0.1064 | [-0.1863, -0.0443] | -0.0121 | 55.4 | 2.51 | 0.0145 |
| TCN | 12 | -0.1 | [-0.1811, -0.0307] | -0.0092 | 53.7 | 1.498 | 0.1388 |
| TimesFM | 1 | -0.0434 | [-0.0632, -0.0279] | -0.0358 | 87.6 | 7.047 | <0.0001 |
| TimesFM | 3 | -0.0268 | [-0.0761, 0.0088] | -0.0153 | 63.5 | 1.209 | 0.2311 |
| TimesFM | 6 | -0.0186 | [-0.0569, 0.0106] | -0.012 | 62 | 0.926 | 0.3577 |
| TimesFM | 12 | 0.0082 | [-0.0198, 0.0289] | 0.0054 | 45.8 | -0.471 | 0.6389 |
| XGBoost | 1 | -0.078 | [-0.1352, -0.0385] | -0.0285 | 80.6 | 4.642 | <0.0001 |
| XGBoost | 3 | -0.1681 | [-0.3006, -0.0744] | -0.07 | 72.9 | 2.808 | 0.00652 |
| XGBoost | 6 | -0.2387 | [-0.4050, -0.1197] | -0.0966 | 70.5 | 2.746 | 0.007746 |
| XGBoost | 12 | -0.2854 | [-0.4680, -0.1553] | -0.1106 | 69.7 | 2.774 | 0.007161 |

**Table S4. Magnitude of Population Dynamics Foundation Model (PDFM) adjustment effects in municipality-month forecasts with high reported dengue case counts.** For each forecasting model and horizon, mean ΔWIS (PDFM-adjusted minus unadjusted WIS) was calculated within the highest positive case-count stratum. High-minus-low contrasts compare mean ΔWIS between the highest and lowest positive case-count strata within each target month. Negative mean ΔWIS indicates improved performance following PDFM adjustment; negative high-minus-low differences indicate greater absolute WIS reductions in the highest case-count stratum. Estimates and 95% confidence intervals were calculated from equally weighted target-month means using a temporal moving-block bootstrap (3-month blocks; 2,000 replicates). Case-count strata were defined separately within each target month and model–horizon combination using the 33rd and 67th percentiles of positive reported case counts. Confidence intervals are unadjusted for multiple comparisons; analyses are exploratory.

| Model | Horizon (months) | High-stratum mean ΔWIS | High-stratum 95% CI | High minus low ΔWIS | High minus low 95% CI |
|---|---|---|---|---|---|
| SARIMA | 1 | -0.072 | [-0.135, -0.015] | -0.059 | [-0.096, -0.016] |
| SARIMA | 3 | -0.28 | [-0.497, -0.091] | -0.373 | [-0.540, -0.219] |
| SARIMA | 6 | -0.283 | [-0.526, -0.061] | -0.416 | [-0.605, -0.228] |
| SARIMA | 12 | -0.091 | [-0.287, 0.050] | -0.221 | [-0.381, -0.094] |
| TCN | 1 | -0.128 | [-0.164, -0.098] | -0.096 | [-0.121, -0.073] |
| TCN | 3 | -0.556 | [-0.687, -0.432] | -0.593 | [-0.705, -0.490] |
| TCN | 6 | -0.77 | [-0.929, -0.614] | -0.846 | [-0.988, -0.689] |
| TCN | 12 | -0.67 | [-0.894, -0.461] | -0.744 | [-0.952, -0.553] |
| XGBoost | 1 | -0.325 | [-0.441, -0.240] | -0.313 | [-0.410, -0.234] |
| XGBoost | 3 | -0.905 | [-1.146, -0.705] | -0.938 | [-1.148, -0.768] |
| XGBoost | 6 | -1.309 | [-1.590, -1.087] | -1.34 | [-1.582, -1.146] |
| XGBoost | 12 | -1.526 | [-1.911, -1.198] | -1.547 | [-1.882, -1.211] |
| N-BEATS | 1 | -2.61 | [-2.881, -2.364] | -2.469 | [-2.687, -2.276] |
| N-BEATS | 3 | -4.573 | [-5.017, -4.192] | -4.424 | [-4.776, -4.081] |

| | | | | | |
|---|---|---|---|---|---|
| N-BEATS | 6 | -5.055 | [-5.520, -4.661] | -4.868 | [-5.211, -4.537] |
| N-BEATS | 12 | -4.865 | [-5.820, -3.953] | -4.669 | [-5.499, -3.717] |
| TimesFM | 1 | -0.043 | [-0.058, -0.024] | 0.032 | [0.014, 0.058] |
| TimesFM | 3 | -0.096 | [-0.152, -0.045] | -0.054 | [-0.081, -0.020] |
| TimesFM | 6 | -0.07 | [-0.109, -0.029] | -0.04 | [-0.059, -0.014] |
| TimesFM | 12 | -0.017 | [-0.043, 0.017] | -0.017 | [-0.039, 0.024] |

**Table S5. Geographical variation in PDFM adjustment effects across Brazil.** Mean changes in Weighted Interval Score (ΔWIS) following Population Dynamics Foundation Model (PDFM) residual adjustment for five forecasting architectures at 1-, 3-, 6- and 12-month horizons across Brazil's five macroregions. Each cell reports mean ΔWIS, with mean log-relative WIS skill in parentheses. ΔWIS was defined as $WIS_{PDFM} - WIS_{unadjusted}$, with negative values indicating improved forecasting performance. Log-relative skill was defined as $\log(1 + WIS_{unadjusted}) - \log(1 + WIS_{PDFM})$, with positive values indicating improvement. Both measures were calculated for individual paired municipality-month forecasts before averaging. Results cover 5,568 municipalities with available PDFM embeddings between January 2020 and August 2025. Nonfinite paired observations were excluded without imputation.

| Geography | Model | Municipalities | 1 month | 3 months | 6 months | 12 months |
|---|---|---|---|---|---|---|
| North | SARIMA | 450 | -0.039 (+0.013) | -0.054 (+0.013) | -0.058 (+0.023) | -0.006 (+0.010) |
| | TCN | | -0.017 (-0.001) | -0.030 (+0.006) | -0.035 (+0.012) | -0.030 (+0.013) |
| | XGBoost | | -0.090 (+0.007) | -0.185 (+0.006) | -0.262 (+0.011) | -0.295 (+0.015) |
| | N-BEATS | | -0.450 | -0.730 | -0.816 | -0.851 |

| | | | | | | |
|---|---|---|---|---|---|---|
| | | | (+0.052) | (+0.065) | (+0.076) | (+0.075) |
| | TimesFM | | -0.036 (-0.006) | -0.026 (-0.016) | -0.020 (-0.016) | -0.007 (-0.024) |
| Northeast | SARIMA | 1793 | -0.043 (+0.019) | -0.043 (+0.013) | -0.014 (+0.010) | -0.008 (+0.022) |
| | TCN | | -0.001 (-0.004) | +0.003 (-0.009) | +0.017 (-0.012) | +0.014 (+0.002) |
| | XGBoost | | -0.025 (-0.002) | -0.049 (-0.021) | -0.073 (-0.034) | -0.082 (-0.035) |
| | N-BEATS | | -0.344 (+0.021) | -0.634 (+0.018) | -0.707 (+0.018) | -0.691 (+0.029) |
| | TimesFM | | -0.037 (-0.005) | -0.014 (-0.019) | -0.013 (-0.018) | -0.003 (-0.024) |
| Central-West | SARIMA | 467 | -0.016 (-0.005) | -0.034 (-0.016) | -0.040 (-0.008) | +0.019 (-0.015) |
| | TCN | | -0.014 (-0.004) | -0.051 (-0.004) | -0.081 (+0.004) | -0.094 (+0.007) |
| | XGBoost | | -0.081 (+0.004) | -0.232 (+0.007) | -0.376 (+0.023) | -0.472 (+0.036) |
| | N-BEATS | | -0.426 (+0.050) | -0.832 (+0.064) | -0.972 (+0.077) | -0.947 (+0.067) |
| | TimesFM | | -0.030 (-0.008) | -0.022 (-0.015) | -0.017 (-0.016) | +0.010 (-0.028) |
| Southeast | SARIMA | 1667 | -0.043 (-0.003) | -0.060 (-0.023) | -0.076 (-0.009) | +0.023 (-0.013) |
| | TCN | | -0.020 (-0.006) | -0.090 (-0.007) | -0.135 (-0.000) | -0.096 (-0.003) |
| | XGBoost | | -0.058 (-0.002) | -0.157 (-0.013) | -0.229 (-0.013) | -0.279 (-0.008) |

| | | | | | | |
|---|---|---|---|---|---|---|
| | N-BEATS | | -0.702 (+0.097) | -1.342 (+0.126) | -1.538 (+0.144) | -1.536 (+0.136) |
| | TimesFM | | -0.039 (-0.008) | -0.022 (-0.024) | -0.011 (-0.024) | +0.041 (-0.039) |
| South | SARIMA | 1191 | -0.113 (+0.002) | -0.181 (-0.013) | -0.173 (-0.014) | -0.082 (-0.026) |
| | TCN | | -0.096 (-0.007) | -0.226 (-0.014) | -0.289 (-0.006) | -0.306 (-0.000) |
| | XGBoost | | -0.181 (+0.003) | -0.331 (-0.010) | -0.440 (-0.001) | -0.524 (+0.012) |
| | N-BEATS | | -0.866 (+0.103) | -1.272 (+0.121) | -1.369 (+0.133) | -1.385 (+0.130) |
| | TimesFM | | -0.067 (-0.003) | -0.055 (-0.024) | -0.038 (-0.024) | -0.016 (-0.025) |

**Table S6. Effect of covariate-set size on forecasting performance.** Values were calculated after averaging ΔWIS across the January 2020–August 2025 target-month evaluation period within each municipality, where $\Delta WIS = WIS_{minimal} - WIS_{full}$. Negative values indicate better performance with the minimal covariate set; positive values indicate better performance with the full covariate set. The full and minimal covariate specifications are defined in Table 1. P25 and P75 denote the 25th and 75th percentiles of municipality-level mean ΔWIS. "Municipalities favouring minimal" is the percentage of municipalities with a negative mean ΔWIS. Models were evaluated across 5,570 Brazilian municipalities at 1-, 3-, 6- and 12-month forecast horizons.

| Model | Horizon (months) | Mean municipality -level ΔWIS | Median municipality -level ΔWIS | P25 | P75 | Municipalities favouring minimal (%) |
|---|---|---|---|---|---|---|
| N-BEATS | 1 | -0.478 | -0.226 | -0.58 | -0.062 | 87.2 |
| N-BEATS | 3 | -0.627 | -0.234 | -0.747 | -0.033 | 80.9 |
| N-BEATS | 6 | -0.647 | -0.234 | -0.767 | -0.029 | 79.7 |
| N-BEATS | 12 | -0.699 | -0.248 | -0.83 | -0.027 | 79.2 |

| | | | | | | |
|---|---|---|---|---|---|---|
| SARIMA | 1 | -0.051 | -0.045 | -0.066 | -0.03 | 98.9 |
| SARIMA | 3 | -0.089 | -0.074 | -0.118 | -0.043 | 98.3 |
| SARIMA | 6 | -0.093 | -0.076 | -0.128 | -0.042 | 97.8 |
| SARIMA | 12 | -0.094 | -0.079 | -0.129 | -0.044 | 98.3 |
| TCN | 1 | 0.034 | 0.041 | -0.001 | 0.093 | 25.3 |
| TCN | 3 | 0.095 | 0.104 | 0 | 0.248 | 24.4 |
| TCN | 6 | 0.141 | 0.129 | 0 | 0.337 | 24.7 |
| TCN | 12 | 0.186 | 0.128 | -0.02 | 0.379 | 26.8 |
| TimesFM | 1 | 0.004 | 0.003 | -0.033 | 0.04 | 47.6 |
| TimesFM | 3 | -0.012 | -0.009 | -0.049 | 0.022 | 59.3 |
| TimesFM | 6 | -0.027 | -0.021 | -0.073 | 0.015 | 65.7 |
| TimesFM | 12 | -0.02 | -0.011 | -0.066 | 0.03 | 59.3 |
| XGBoost | 1 | -0.018 | -0.017 | -0.063 | 0.022 | 60.3 |
| XGBoost | 3 | 0.018 | 0 | -0.083 | 0.107 | 48.5 |
| XGBoost | 6 | 0.078 | 0.015 | -0.078 | 0.206 | 42.5 |
| XGBoost | 12 | 0.167 | 0.048 | -0.056 | 0.32 | 37 |

**Table S7. Impact of independent municipality-level versus joint state-grouped fitting.** For each model and forecast horizon, $\Delta WIS = WIS_{independent} - WIS_{state\text{-}grouped}$ was averaged across the January 2020–August 2025 target-month evaluation period within each of the 20 municipalities with the highest cumulative reported dengue case counts over the study period, then summarised across municipalities. Negative values favour univariate fitting; positive values favour multivariate fitting. P25 and P75 describe the distribution of municipality-level mean ΔWIS. "Univariate better" is the percentage of municipalities with a negative mean ΔWIS. Two-sided Wilcoxon signed-rank tests were applied to the 20 municipality-level values.

| Model | Horizon | n | Mean ΔWIS | Median ΔWIS | P25 | P75 | Independent fitting better (%) | W | p-value |
|---|---|---|---|---|---|---|---|---|---|

| | | | | | | | | | |
|---|---|---|---|---|---|---|---|---|---|
| N-BEATS | 1 | 20 | -2.828 | -3.581 | -5.217 | -1.683 | 85 | 27 | 0.0023 |
| N-BEATS | 3 | 20 | -6.853 | -7.989 | -10.182 | -5.286 | 90 | 10 | <0.0001 |
| N-BEATS | 6 | 20 | -8.213 | -9.29 | -11.634 | -5.997 | 90 | 7 | <0.0001 |
| N-BEATS | 12 | 20 | -9.843 | -10.567 | -13.324 | -8.266 | 90 | 4 | <0.0001 |
| SARIMA | 1 | 20 | -0.002 | -0.002 | -0.005 | 0.001 | 70 | 60 | 0.0973 |
| SARIMA | 3 | 20 | -0.007 | -0.006 | -0.012 | -0.002 | 75 | 45 | 0.024 |
| SARIMA | 6 | 20 | -0.002 | 0.001 | -0.006 | 0.008 | 50 | 101 | 0.8983 |
| SARIMA | 12 | 20 | -0.003 | -0.005 | -0.011 | 0.003 | 65 | 67 | 0.165 |
| TCN | 1 | 20 | 1.157 | 0.82 | 0.642 | 1.291 | 0 | 0 | <0.0001 |
| TCN | 3 | 20 | 0.58 | 0.051 | -0.193 | 0.368 | 45 | 91 | 0.6215 |
| TCN | 6 | 20 | 0.436 | 0.111 | -0.393 | 0.225 | 35 | 104 | 0.9854 |
| TCN | 12 | 20 | 0.589 | 0.006 | -0.324 | 0.205 | 50 | 102 | 0.9273 |
| TimesFM | 1 | 20 | 0.036 | 0.044 | 0.006 | 0.057 | 25 | 20 | 0.0007 |
| TimesFM | 3 | 20 | 0.144 | 0.14 | 0.093 | 0.186 | 5 | 1 | <0.0001 |
| TimesFM | 6 | 20 | 0.178 | 0.148 | 0.106 | 0.242 | 0 | 0 | <0.0001 |
| TimesFM | 12 | 20 | 0.254 | 0.218 | 0.143 | 0.337 | 0 | 0 | <0.0001 |
| XGBoost | 1 | 20 | 0.636 | 0.326 | -0.233 | 0.938 | 35 | 53 | 0.0532 |
| XGBoost | 3 | 20 | -0.028 | -0.019 | -0.775 | 0.326 | 55 | 93 | 0.6742 |
| XGBoost | 6 | 20 | -0.784 | -0.78 | -1.607 | -0.122 | 75 | 34 | 0.0064 |
| XGBoost | 12 | 20 | -1.329 | -1.1 | -2.283 | -0.375 | 90 | 13 | 0.0002 |

**Table S8. Impact of fitting to aggregated case series versus aggregating municipality-level probabilistic forecasts.** For each model and forecast horizon, ΔWIS was defined as WIS for forecasts fitted directly to aggregated state-level case series minus WIS for aggregated municipality-level probabilistic forecasts. ΔWIS was averaged across the January 2020–August 2025 target-month evaluation period within each of the 27 Brazilian states, then summarised across states. Negative values favour direct fitting to aggregated case series; positive values favour aggregation of municipality-level forecasts. P25 and P75 describe the distribution of state-level mean ΔWIS. "States favouring direct aggregate fitting" is the percentage of states with a negative mean ΔWIS. Two-sided Wilcoxon signed-rank tests were applied to the 27 state-level values.

| Model | Horizon | n | Mean ΔWIS | Median ΔWIS | P25 | P75 | States favouring direct aggregate fitting | W | p-value |
|---|---|---|---|---|---|---|---|---|---|

| | | | | | | | | | |
|---|---|---|---|---|---|---|---|---|---|
| | | | | | | | (%) | | |
| N-BEATS | 1 | 27 | -0.671 | -0.314 | -2.626 | 1.003 | 59.3 | 140 | 0.2483 |
| N-BEATS | 3 | 27 | -3.217 | -3.15 | -5.615 | -0.596 | 81.5 | 46 | 0.0003 |
| N-BEATS | 6 | 27 | -4.12 | -4.086 | -6.874 | -1.539 | 81.5 | 40 | 0.0001 |
| N-BEATS | 12 | 27 | -4.955 | -5.296 | -7.668 | -1.814 | 77.8 | 37 | <0.0001 |
| SARIMA | 1 | 27 | 0.006 | 0.102 | 0.024 | 0.148 | 22.2 | 89 | 0.0151 |
| SARIMA | 3 | 27 | -0.185 | 0.045 | -0.274 | 0.143 | 44.4 | 167 | 0.6109 |
| SARIMA | 6 | 27 | -0.24 | -0.065 | -0.365 | 0.178 | 51.9 | 159 | 0.4846 |
| SARIMA | 12 | 27 | 0.012 | -0.02 | -0.422 | 0.508 | 55.6 | 181 | 0.8593 |
| TCN | 1 | 27 | 1.22 | 0.785 | 0.729 | 1.392 | 0 | 0 | <0.0001 |
| TCN | 3 | 27 | 0.944 | 0.689 | 0.452 | 1.117 | 0 | 0 | <0.0001 |
| TCN | 6 | 27 | 1.123 | 1.011 | 0.674 | 1.471 | 0 | 0 | <0.0001 |
| TCN | 12 | 27 | 0.914 | 0.899 | 0.48 | 1.299 | 7.4 | 17 | <0.0001 |
| TimesFM | 1 | 27 | 0.089 | 0.168 | 0.076 | 0.27 | 14.8 | 56 | 0.0008 |
| TimesFM | 3 | 27 | 0.164 | 0.238 | 0.192 | 0.357 | 11.1 | 56 | 0.0008 |
| TimesFM | 6 | 27 | 0.245 | 0.281 | 0.209 | 0.392 | 7.4 | 29 | <0.0001 |
| TimesFM | 12 | 27 | 0.236 | 0.225 | 0.18 | 0.421 | 7.4 | 29 | <0.0001 |
| XGBoost | 1 | 27 | 1.049 | 0.781 | 0.451 | 1.273 | 0 | 0 | <0.0001 |
| XGBoost | 3 | 27 | 0.272 | 0.336 | -0.121 | 0.618 | 37 | 110 | 0.0585 |
| XGBoost | 6 | 27 | -0.48 | -0.487 | -0.937 | 0.263 | 66.7 | 75 | 0.005 |
| XGBoost | 12 | 27 | -1.113 | -1.162 | -1.93 | -0.208 | 81.5 | 30 | <0.0001 |

**Table S9. Impact of monthly iterative model retraining in Ceará and São Paulo.** Models were compared under the initial training regime and monthly iterative retraining. For each state, model and forecast horizon, WIS was averaged across the January 2020–August 2025 target-month evaluation period within each municipality, and the resulting municipality-level means were compared using two-sided Wilcoxon signed-rank tests. Median difference was defined as initial-training WIS minus monthly-retraining WIS; negative values therefore indicate better performance under initial training. Ceará comparisons included 184 municipalities and São Paulo comparisons included 645 municipalities. Complete comparisons were available for SARIMA and XGBoost in both states and for TCN in Ceará. For N-BEATS, non-finite or otherwise unusable WIS summaries prevented valid paired comparisons in São Paulo and Ceará; N-BEATS was therefore excluded from the retraining performance comparison.

TimesFM was excluded because its pretrained model parameters were not fitted or fine-tuned to the study data, making iterative retraining inapplicable.

| State | Model | Horizon | n | W statistic | p-value | Median difference |
|---|---|---|---|---|---|---|
| Ceará | SARIMA | 1 | 184 | 8266 | 0.7359 | 0.0004 |
| Ceará | SARIMA | 3 | 184 | 7585 | 0.201 | 0.0015 |
| Ceará | SARIMA | 6 | 184 | 8121 | 0.5908 | -0.0009 |
| Ceará | SARIMA | 12 | 184 | 7857 | 0.3667 | -0.0017 |
| Ceará | TCN | 1 | 184 | 0 | <0.0001 | -2.0236 |
| Ceará | TCN | 3 | 184 | 0 | <0.0001 | -1.647 |
| Ceará | TCN | 6 | 184 | 15 | <0.0001 | -1.5385 |
| Ceará | TCN | 12 | 184 | 24 | <0.0001 | -1.838 |
| Ceará | XGBoost | 1 | 184 | 184 | <0.0001 | -0.4133 |
| Ceará | XGBoost | 3 | 184 | 1173 | <0.0001 | -0.2596 |
| Ceará | XGBoost | 6 | 184 | 3251 | <0.0001 | -0.2491 |
| Ceará | XGBoost | 12 | 184 | 3676 | <0.0001 | -0.2541 |
| São Paulo | SARIMA | 1 | 645 | 98151 | 0.2038 | 0.0003 |
| São Paulo | SARIMA | 3 | 645 | 100215 | 0.4038 | -0.0004 |
| São Paulo | SARIMA | 6 | 645 | 98352 | 0.2193 | -0.0003 |
| São Paulo | SARIMA | 12 | 645 | 100404 | 0.4266 | 0.0017 |
| São Paulo | XGBoost | 1 | 645 | 1294 | <0.0001 | -0.7012 |
| São Paulo | XGBoost | 3 | 645 | 7176 | <0.0001 | -0.5147 |
| São Paulo | XGBoost | 6 | 645 | 24297 | <0.0001 | -0.4512 |
| São Paulo | XGBoost | 12 | 645 | 10617 | <0.0001 | -0.7844 |

**Table S10. Out-of-sample probabilistic forecasting performance during the independent September 2023–August 2025 hold-out period.** Forecasts from the independently developed ERA model and all five primary forecasting models were evaluated across 5,570 Brazilian municipalities, comprising 133,680 municipality-month forecasts at each forecast horizon. Values summarise WIS across municipality-month forecasts; lower values indicate better probabilistic forecast performance. For this hold-out comparison, WIS was calculated on the original case-count scale and is therefore not directly comparable with WIS values from the primary log(1 + cases)-scale evaluation. Median and upper quantiles are additionally reported because WIS distributions were strongly right-skewed. The extreme long-horizon mean WIS values for SARIMA reflect a small number of exceptionally large forecast errors.

| Model | Horizon (months) | Mean WIS | Median WIS | P90 WIS | P95 WIS |
|---|---|---|---|---|---|
| ERA | 1 | 296.1 | 5.97 | 284.3 | 766.1 |
| ERA | 3 | 335.9 | 6.53 | 325.6 | 871.7 |
| ERA | 6 | 357.8 | 6.28 | 347.9 | 926 |
| ERA | 12 | 384 | 6.6 | 384.7 | 1021.4 |
| SARIMA | 1 | 154.9 | 6.04 | 193.5 | 478.2 |
| SARIMA | 3 | 348.9 | 9.43 | 384.9 | 974.7 |
| SARIMA | 6 | 7731.6 | 10.77 | 513.4 | 1304 |
| SARIMA | 12 | $7.208 \times 10^9$ | 10.14 | 690.3 | 2072 |
| TCN | 1 | 156.3 | 5.81 | 151.5 | 375.5 |
| TCN | 3 | 290.4 | 7.62 | 254.2 | 679.1 |
| TCN | 6 | 325.2 | 8.13 | 304.4 | 810 |
| TCN | 12 | 367 | 8.45 | 324.6 | 870.3 |
| N-BEATS | 1 | 580.8 | 4.54 | 331.2 | 982.4 |
| N-BEATS | 3 | 511.2 | 4.89 | 441.1 | 1246.9 |
| N-BEATS | 6 | 506.2 | 5.17 | 461.7 | 1288.9 |
| N-BEATS | 12 | 498.6 | 5.61 | 479.3 | 1312.9 |
| TimesFM | 1 | 2416.7 | 6.36 | 198.5 | 490.9 |
| TimesFM | 3 | 326.9 | 5.83 | 221.8 | 553.8 |
| TimesFM | 6 | 164.1 | 6.48 | 204.3 | 505.4 |
| TimesFM | 12 | 173.5 | 6.2 | 207.8 | 505.6 |
| XGBoost | 1 | 291.3 | 5.24 | 159.1 | 477.8 |
| XGBoost | 3 | 366.2 | 6.08 | 264.4 | 807.8 |

| | | | | | |
|---|---|---|---|---|---|
| XGBoost | 6 | 387.9 | 6.08 | 306.9 | 904.6 |
| XGBoost | 12 | 394.7 | 6.04 | 322.6 | 928.8 |

**Table S11. Information sources and model components incorporated into the final ERA-derived dengue forecasting model.** The table summarises the information sources selected by ERA and their roles in the resulting forecasting model.

| Information source | Component | Role in ERA-derived model |
|---|---|---|
| Epidemiological | Historical reported dengue cases | Municipality-specific seasonal baseline |
| Epidemiological | Recent reported dengue cases | Recent epidemic dynamics |
| Spatial | Municipality adjacency | Spatial propagation of recent epidemic dynamics |
| Demographic | Population | Population-dependent additive contribution to expected case counts |
| Climatic | ONI | Large-scale climatic effects |
| Climatic | SPI-6 | Precipitation and drought-related climatic effects |
| Contextual | PDFM embeddings | Municipality-specific model parameterisation |
| Temporal | Forecast horizon | Horizon-dependent attenuation of recent epidemic dynamics |
| Probabilistic | Negative Binomial distribution | Generation of probabilistic forecasts |

**Supplementary Table S12. Summary of modelling findings and implications for probabilistic dengue forecasting in Brazil.** Synthesis of the principal findings from comparisons of five forecasting architectures (SARIMA, TCN, XGBoost, N-BEATS and TimesFM), forecast horizons, geographical settings, Population Dynamics Foundation Model (PDFM) residual adjustment, covariate selection, spatial fitting, forecast aggregation and model retraining. Forecasting performance was assessed primarily using the Weighted Interval Score (WIS), with lower values indicating better probabilistic forecasts. Findings reflect the specific architectures, configurations and retrospective evaluation settings examined in this study and should not be interpreted as identifying universally optimal approaches for dengue forecasting in Brazil. Spatial fitting comparisons were restricted to 20 high-burden municipalities, and monthly retraining was evaluated only in Ceará and São Paulo.

| Modelling consideration | Findings from this study | Implications for forecasting practice |
|---|---|---|
| Forecasting architecture and horizon | Among the five architectures evaluated, TCN achieved the lowest mean WIS at one month, while TimesFM performed best at 3-, 6- and 12-month horizons. | TCN and TimesFM warrant consideration for short- and longer-horizon forecasting, respectively, alongside other established dengue forecasting approaches. |
| Geographical variation | Model rankings were consistent across Brazil's five macroregions, although the magnitude of performance differences varied. TCN's one-month advantage over TimesFM was substantially weaker in Rio Grande do Sul and Santa Catarina. | National performance rankings may inform model selection, but geographical differences should be considered when evaluating approaches for particular locations. |
| PDFM residual adjustment | PDFM adjustment improved mean WIS in 19 of 20 model–horizon combinations, with the largest improvements for N-BEATS and XGBoost. Benefits varied by architecture, horizon and geographical setting. | PDFM embeddings may provide additional predictive information, but larger adjustment benefits do not necessarily imply better overall forecasting performance. Adjusted models should be evaluated against strong unadjusted alternatives. |
| Covariate selection | Minimal covariates generally favoured SARIMA and N-BEATS, while TCN benefited from the full covariate set. XGBoost showed horizon-dependent effects, and TimesFM was comparatively insensitive to covariate selection. | Additional covariates should not be assumed to improve performance. Covariate selection should account for forecasting architecture and horizon. |
| Spatial fitting strategy | Independent municipality-level fitting favoured N-BEATS, whereas state-grouped fitting favoured TimesFM. TCN favoured state-grouped fitting at one month, while XGBoost favoured independent fitting at longer | Within the evaluated high-burden municipalities, independent fitting was generally preferable for N-BEATS, while state-grouped fitting favoured TimesFM. These findings should be validated before extending recommendations to other |

| | | |
|---|---|---|
| | horizons. Comparisons involved 20 high-burden municipalities. | municipalities. |
| State-level forecast aggregation | Aggregating municipality-level probabilistic forecasts generally favoured TCN and TimesFM, whereas direct fitting to aggregated state-level case series favoured N-BEATS at longer horizons. XGBoost exhibited horizon-dependent differences. | The approach used to generate state-level forecasts should be considered part of model design, with the preferred strategy depending on architecture and forecast horizon. |
| Model retraining | Monthly retraining provided no consistent improvement in Ceará and São Paulo. XGBoost favoured initial training, while N-BEATS retraining encountered numerical instability. | Frequent retraining should not be assumed to improve forecasting performance. Updating schedules should be evaluated against predictive accuracy, computational requirements and model stability. |

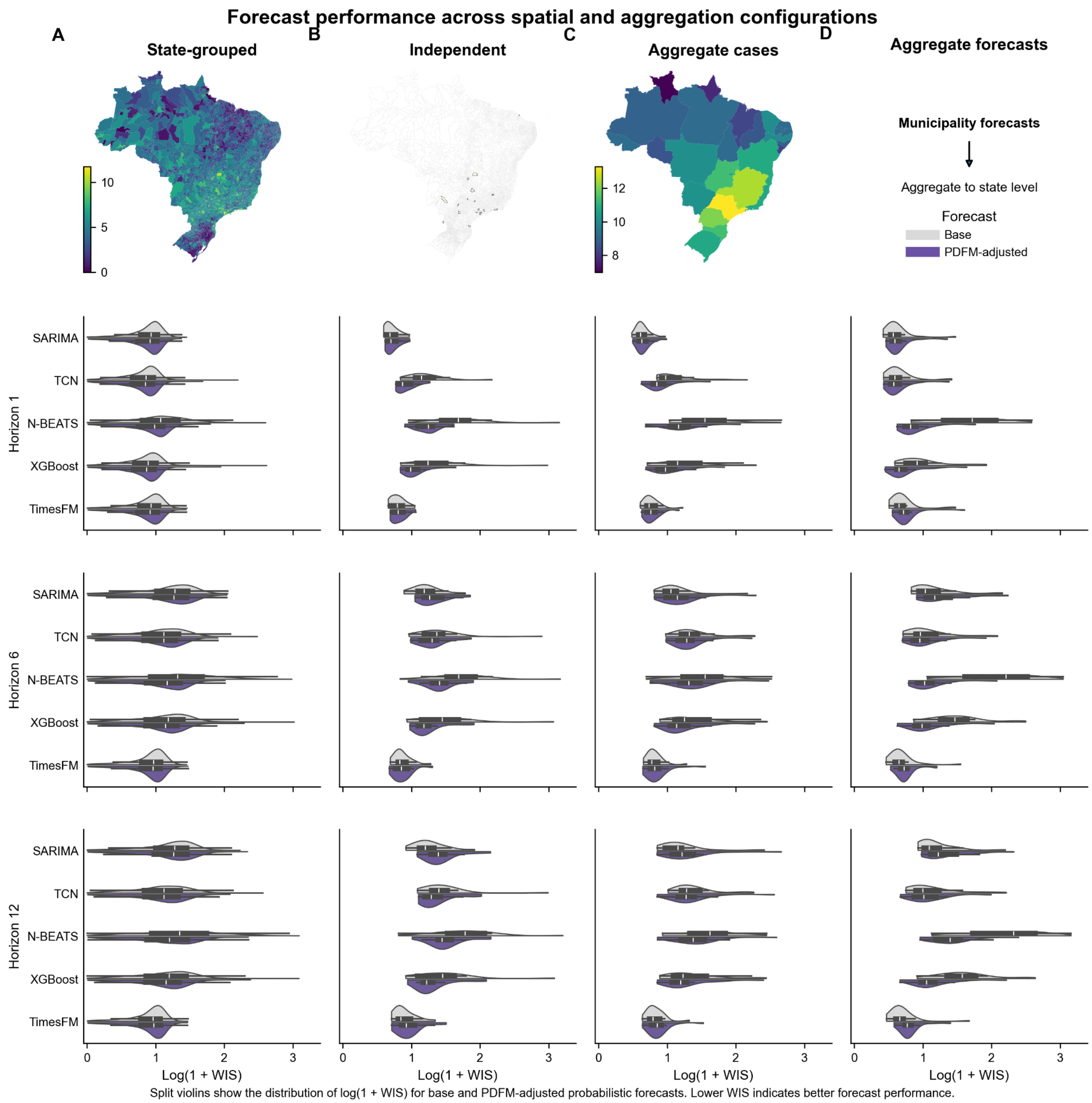


**Figure S1. Distributional effects of forecasting configuration on WIS at 1-, 6- and 12-month horizons.** State-grouped forecasts are obtained from models fitted jointly across municipalities within each state, whereas independent forecasts are obtained from models fitted separately to individual municipalities. Both configurations are evaluated using municipality-level forecasts for the same 20 municipalities with the highest cumulative reported dengue case counts over the study period, highlighted on the map. Aggregate cases denote forecasts from models fitted directly to state-level aggregated case series, whereas aggregate forecasts denote municipality-level probabilistic forecasts aggregated to state level. For PDFM adjustment of models fitted directly to aggregated state-level case series, state-level PDFM representations were constructed by averaging municipality-level embeddings within each state. Split violins show the distribution of log(1 + mean WIS) across spatial units, where mean WIS was calculated over the January 2020–August 2025 evaluation period within each municipality or state. Lower values indicate better probabilistic forecast performance.

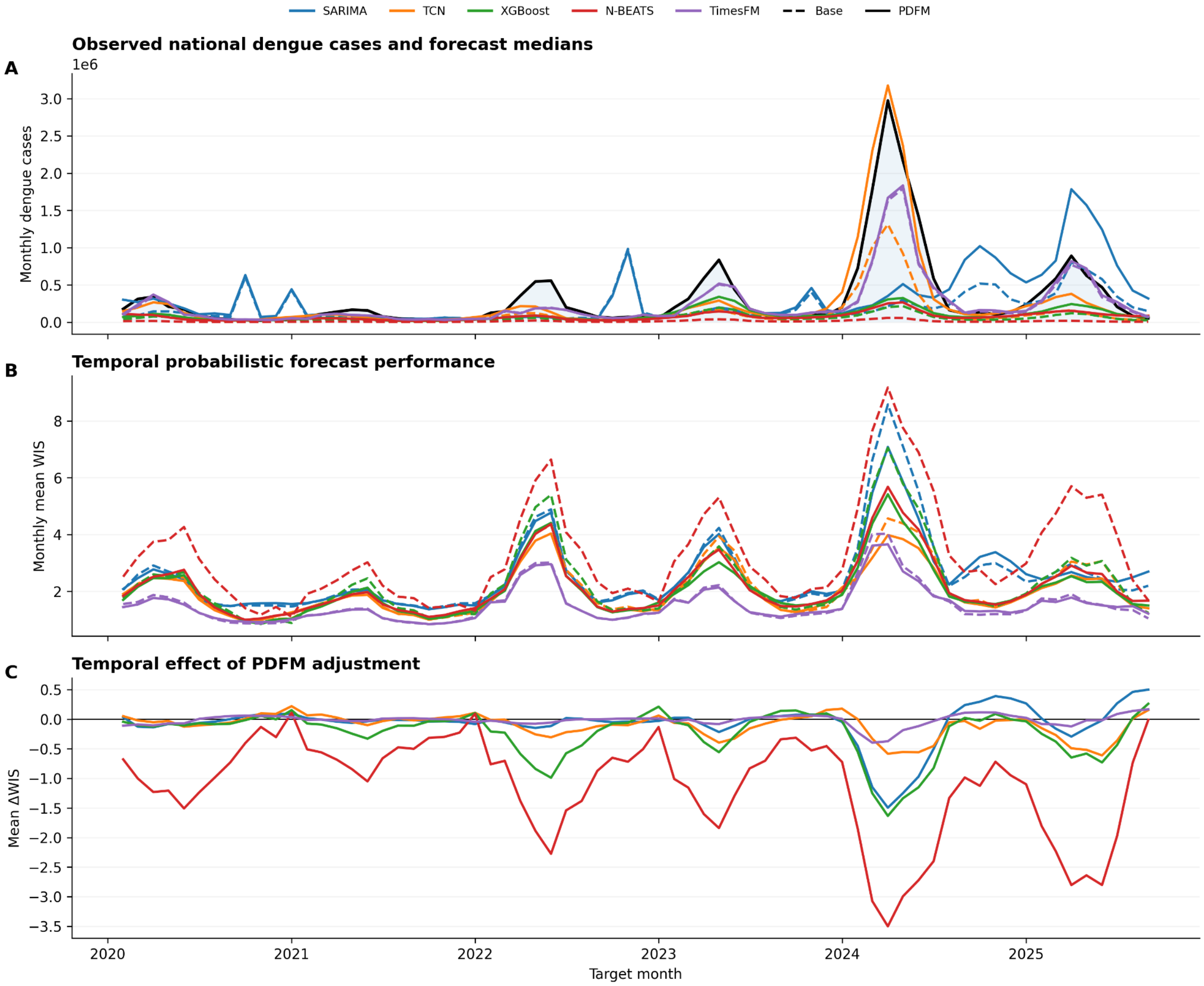


**Figure S2. Temporal forecast performance and effects of Population Dynamics Foundation Model (PDFM) adjustment at the 6-month forecast horizon.** (A) Observed national monthly dengue cases and national aggregates of municipality-level median forecasts for each forecasting model before PDFM adjustment (Base; dashed lines) and after PDFM residual adjustment (PDFM; solid lines). National forecast series were constructed by summing municipality-level median forecasts. Individual municipality-level median predictions exceeding 1 million cases were treated as missing for this panel only to prevent extreme predictions from obscuring the displayed forecast trajectories; these forecasts were retained in the quantitative WIS analyses. (B) Monthly mean Weighted Interval Score (WIS) across municipalities for unadjusted and PDFM-adjusted forecasts. (C) Monthly mean change in WIS following PDFM adjustment ($\Delta WIS = WIS_{PDFM} - WIS_{base}$, where base denotes the corresponding unadjusted forecast). Negative values indicate improved probabilistic forecast performance following PDFM adjustment. Dates denote forecast target months.

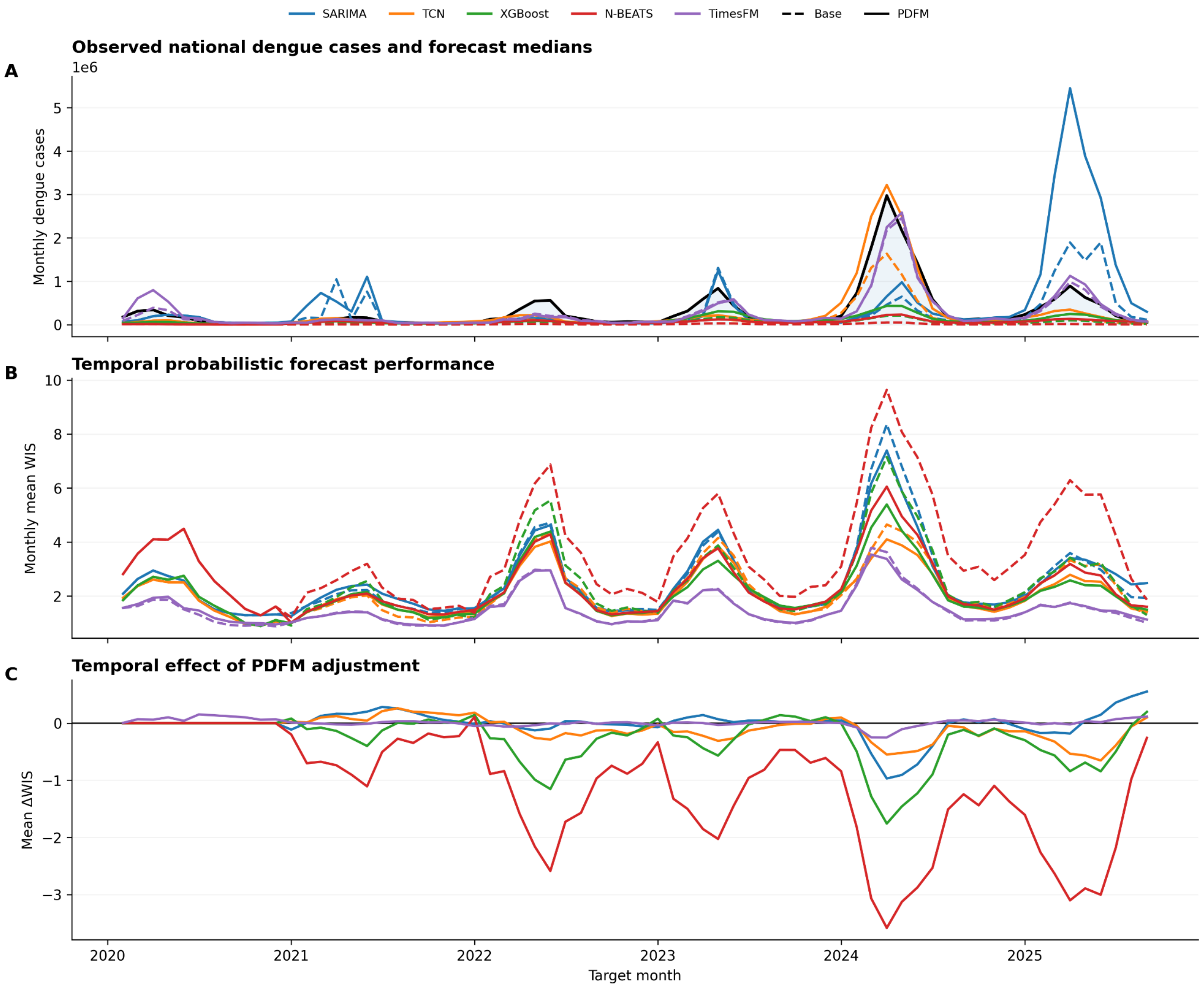


**Figure S3. Temporal forecast performance and effects of Population Dynamics Foundation Model (PDFM) adjustment at the 12-month forecast horizon.** (A) Observed national monthly dengue cases and national aggregates of municipality-level median forecasts for each forecasting model before PDFM adjustment (Base; dashed lines) and after PDFM residual adjustment (PDFM; solid lines). National forecast series were constructed by summing municipality-level median forecasts. Individual municipality-level median predictions exceeding 1 million cases were treated as missing for this panel only to prevent extreme predictions from obscuring the displayed forecast trajectories; these forecasts were retained in the quantitative WIS analyses. (B) Monthly mean Weighted Interval Score (WIS) across municipalities for unadjusted and PDFM-adjusted forecasts. (C) Monthly mean change in WIS following PDFM adjustment ($\Delta WIS = WIS_{PDFM} - WIS_{base}$, where base denotes the corresponding unadjusted forecast). Negative values indicate improved probabilistic forecast performance following PDFM adjustment. Dates denote forecast target months.
